# Machine learning differentiation of Parkinson’s disease and normal pressure hydrocephalus using wearable sensors capturing gait impairments

**DOI:** 10.1101/2025.01.07.24318198

**Authors:** Stefano Magni, Rene Peter Bremm, Konstantinos Verros, Xin He, Sylvie Lecossois, Finn Jelke, Andreas Husch, Jorge Gonçalves, Frank Hertel

## Abstract

Gait impairments in patients with Parkinson’s Disease (PD) and Normal Pressure Hydrocephalus (NPH) are diagnosed with visual clinical assessments. Despite standardized gait tests and clinicians’ expertise, such approaches can be subjective and challenging due to similar symptoms between the two diseases. Wearable sensors and machine learning (ML) can assist clinicians by offering objective and quantitative assessments of gait impairments that can help distinguishing between PD and NPH. This study consists of a cohort of 12 PD and 11 NPH patients that performed standardized gait tests. Gait was measured by wearable sensors embedded in patients’ shoes: a three-axis gyroscope, a three-axis accelerometer and eight pressure sensors in each insole. Sensors and computational pipeline to extract gait cycle features were validated and calibrated on 21 healthy subjects. ML approaches were employed to identify changes in gait cycle features between the PD and NPH patients groups. Twenty-seven ML classifiers were compared, leading to select linear support vector machines, resulting in a classification accuracy of 0.70 ± 0.28 and an area under the ROC curve of 0.74 ± 0.39. Combining wearable sensors with ML algorithms trained on gait cycle features from those sensors showed the potential for objective differentiation of gait patterns between PD and NPH patients.

## 1 Introduction

Parkinson’s Disease (PD) leads to debilitating motor symptoms, including tremor, stiffness, bradykinesia, and postural instability [1]. Gait abnormalities in PD are characterized by shortened step and stride length and gait velocity. PD also includes non-motor symptoms, such as cognitive impairments. Another neurological disorder, Normal Pressure Hydrocephalus (NPH), also leads to walking difficulties and cognitive impairments [2]. The gait abnormalities in NPH are characterized by outwardly rotated feet and a reduced step height and length. Both diseases share similar gait impairments and are difficult to distinguish from one another in the early disease stages. However, they require different treatments, so an exact and timely diagnosis is important for improving the long-term outcome.

The severity of motor symptoms is typically assessed via the UPDRS-III scale for PD [3]. There is no unique scale for the assessment of gait impairments in NPH, however a variety of scales could be used [4, 5, 6]. While these standardized assessments provide semi-quantitative measurements of gait impairments, their estimations are based on subjective observations by clinical experts. Thus, there is a strong need for more objective quantitative measurement tools to aid the diagnosis.

Wearable devices equipped with various types of sensors, e.g. accelerometers and gyroscopes, often embedded in inertial measurement units (IMUs), are widely used in gait research in both healthy adults [7] and people with movement disorders [8, 9, 10]. These devices can detect abnormal gait patterns, such as freezing episodes in patients with PD [11, 12]. They have already been used to complement clinical evaluations for NPH assessment and prognosis [6]. IMUs are also used for detecting walking speed, step counting [13], and fall detection [14]. Pressure sensors embedded in shoe insoles have been used for gait monitoring [15] and for the detection of freezing of gait [16]. They have been applied to characterize different walking patterns in healthy subjects [17], and to monitor gait changes upon treatment in NPH [18]. However, the gold standard for gait analysis in movement disorders is actually the gait lab, where extended analysis of gait patterns are performed on a treadmill. This equipment is expensive and does not mimic real life conditions.

The field of Artificial Intelligence is quickly advancing and receiving a lot of attention, and machine learning (ML) algorithms are increasingly employed in research for biomedical and clinical applications. In combination with data from wearable sensors, feature extraction methods and ML algorithms have been used e.g. for fall detection [19] and, in the context of PD, for freezing of gait prediction [20]. They have also been applied to data from pressure sensors embedded in shoe insoles to diagnose PD using public datasets [21, 22], and to classify PD’s medication states and severity levels [23]. The rise of ML and digital technologies to monitor PD is discussed in [24]. ML has also been applied to MRI images of NPH patients for detection of the disease using voxel-based morphometry [25], and for the prediction of response to treatment [26, 27]. Deep Learning (DL) has recently become extremely popular, and applications include early detection of PD via gait analysis from 3D video-based pose estimation in real time [28], activity recognition from multimodal data from wearable sensors [29], and quantitative gait analysis of NPH from video recordings [30]. A comparison of the performances of ML with feature extraction and DL to classify various physical activities from IMUs can be found in [31].

Patients often have to be hospitalized for a significant amount of time to perform the differential diagnosis of different movement disorders via clinical tests and observations. Furthermore, adaptations of treatments in PD patients, such as Deep Brain Stimulation (DBS) parameters or medical treatments, have to be based on long-term clinical observations under hospital conditions. Also, the decision to implant a Cerebrospinal Fluid (CSF) drainage in NPH patients is commonly based on the result of a temporary CSF release (Spinal Tap Test) and the subjective analysis of its effects. Therefore, this study addresses the following research questions. 1) Is it possible to use wearable sensors (IMU, pressure sensors) to get accurate results to diagnose PD versus NPH without using a traditional gait lab? 2) Is it possible to develop and introduce ML-based methods for the analysis of those sensor’s signals? 3) Is it possible with these wearable sensors and the developed analysis methods to differentiate PD from NPH?

These can be summarized in the aim of this study, which is to answer the research question of whether it is possible to distinguish PD from NPH by using ML algorithms trained with gait data from wearable motion sensors, hence aiding in a more objective diagnosis in the future. This would ease the process for measuring gait impairments and could be complementary to expensive treadmills which are the current gold standard for gait assessment.

This paper develops a computational pipeline to extract gait cycle parameters from wearable sensors including accelerometer, gyroscope, and foot pressure sensors embedded in the shoes of 12 PD and 11 NPH patients. The pipeline was then used for statistical analysis and to provide the features necessary to train ML algorithms to distinguish between gait impairments of patients with PD and NPH. This work represents a feasibility study providing a proof-of-concept for a future differential diagnosis of PD versus NPH based on wearable sensors and ML.

## 2 Results

### 2.1 Overview of study, cohort and extracted features

The proof-of-concept study presented in this manuscript is outlined in Fig. 1. First, healthy subjects were considered to validate the wearable sensors (IEE’s ActiSense device) and computational pipeline (Fig. 1A, B). Then, the main clinical study on PD and NPH patients (Fig. 1C) addressed the research questions of this work. Details are provided in the Methods (in Sec. 4.1 and in the following sections) and in the supplementary material.

**Figure 1:**
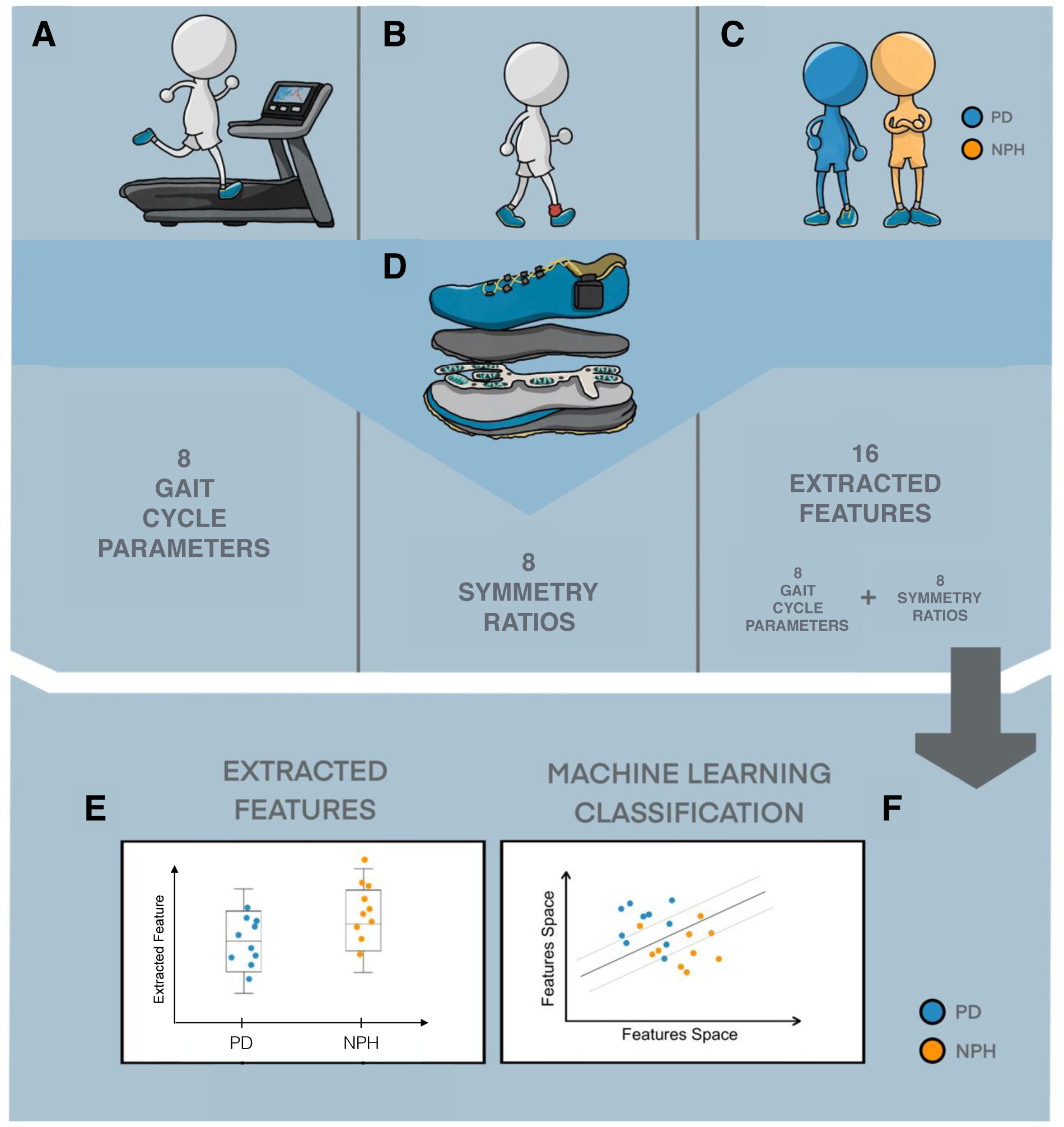
Study overview. **(A)** System validation study (SVS) on healthy subjects using a treadmill. **(B)** Asymmetry validation study (AVS) on healthy subjects employing weights to induce gait asymmetry. **(C)** Clinical study (CS), involving Parkinson’s Pisease (PD) and Normal Pressure Hydrocephalus (NPH) patients. **(D)** Wearable sensors used. **(E)** Statistical analysis of the data to identify significant differences between the gait patterns of PD and NPH patients. **(F)** Machine learning classifiers trained on the data in order to distinguish PD from NPH patients.

All the subjects (Table 1) were equipped with prototype wearable sensors (Fig. 1D and Sec. 4.2) embedded in standardized shoes and were asked to perform standardized walking tests. The gathered raw time-series data were then processed (Methods Sec. 4.3) by a computational pipeline developed in this study, to extract eight gait cycle parameters and eight symmetry ratios, for a total of 16 extracted features summarized in Table 2.

**Table 1:** Demographics and clinical characteristics of study participants. SVS stands for system validation study, AVS for asymmetry validation study, and CS for clinical study.

| Biometric | Healthy (SVS) | Healthy (AVS) | PD (CS) | NPH (CS) |
| --- | --- | --- | --- | --- |
| Number | 9 | 12 | 12 | 11 |
| Sex (m/f) | 8/1 | 6/6 | 9/3 | 6/5 |
| Age (years) | $28.0 \pm 5.0$ | $32.7 \pm 8.4$ | $62.2 \pm 9.3$ | $74.1 \pm 7.1$ |
| Shoes Size [EU] | 39 – 45 | 37 – 45 | 40 – 44 | 37 – 44 |
| Height (cm) | $181.22 \pm 7.33$ | $172.08 \pm 13.5$ | — | — |
| Body mass (kg) | $80.04 \pm 13.19$ | $67.91 \pm 15.54$ | — | — |

**Table 2:** Extracted features. Lines 1-8: gait cycle parameters. Lines 9-16: symmetry ratios of the gait cycle parameters. The complete definition of each feature can be found in Methods, Table 3.

| Extracted Feature | Unit of measure |
| --- | --- |
| Mean gait cycle duration, right ($GCD_R$) | s |
| Mean stance phase, right ($STP_R$) | Percentage of $GCD_R$ |
| Mean swing phase, right ($SWP_R$) | Percentage of $GCD_R$ |
| Cadence, right ($C_R$) | $\frac{\text{steps}}{\text{min}}$ |
| Mean gait cycle duration, left ($GCD_L$) | s |
| Mean stance phase, left ($STP_L$) | Percentage of $GCD_L$ |
| Mean swing phase, left ($SWP_L$) | Percentage of $GCD_L$ |
| Cadence, left ($C_L$) | $\frac{\text{steps}}{\text{min}}$ |
| Symmetry ratio R/L of GCD | adimensional |
| Symmetry ratio R/L of STP | adimensional |
| Symmetry ratio R/L of SWP | adimensional |
| Symmetry ratio R/L of C | adimensional |
| Symmetry ratio Max/Min of GCD | adimensional |
| Symmetry ratio Max/Min of STP | adimensional |
| Symmetry ratio Max/Min of SWP | adimensional |
| Symmetry ratio Max/Min of C | adimensional |

A “system validation study” (SVS, Fig. 1A and Methods Sec. 4.4), was performed on healthy subjects to calibrate and validate the ensemble of wearable sensors and computational pipeline against an instrumented treadmill with integrated force sensors, representing the state-of-the-art in clinical gait assessment. Here, the aim is to assess how precisesely the ActiSense device and pipeline measure gait cycle parameters, and this validated the eight extracted gait cycle parameters (first half of Table 2).

An “asymmetry validation study” (AVS, Fig. 1B and Methods Sec. 4.5) was then performed to assess and validate the capability of the ActiSense device and the developed computational pipeline to measure gait cycle asymmetry on healthy subjects. This validated the extracted eight symmetry ratios (second half of Table 2). This also had the goal of characterizing the magnitude of asymmetry affecting the gait of healthy subjects, providing a baseline for further assessment of asymmetry in diseased subjects. In total, 21 healthy participants (Table 1) took part in these two validation studies.

The main study comparing PD and NPH patients is referred to as “clinical study” (CS), Fig. 1C, and the cohort used therein consisted of 12 PD and 11 NPH patients. Gait data from this cohort were collected from patients hospitalized in the National Department of Neurosurgery at the Centre Hospitalier du Luxembourg. Each patient performed a standardized test consisting of a 10 m walk [32], equipped with the wearable sensors (Fig. 1D and Methods Sec. 4.2), repeating the test twice. These data were used to compute gait cycle parameters and symmetry ratios, then employed to distinguish PD and NPH by statistical data analysis (Fig. 1E and Sec. 2.2) and as features to train ML models (Fig. 1F and Sec. 2.3).

### 2.2 Statistical comparison of extracted features between PD and NPH

For each of the 16 extracted features (Table 2), a statistical comparison was performed to assess if there were significant differences between the measurements performed on the PD and NPH groups (Methods Sec. 4.7). The following eight features are significantly different between the PD and NPH groups: mean stance phase R, mean swing phase R, mean gait cycle duration R and L, cadence R and L, symmetry ratio R/L of stance phase, and symmetry ratio R/L of swing phase (Fig. 2; the complete statistical comparison is in Fig. S1). The features that are significantly different for both the right and left sides between PD and NPH include mean gait cycle duration and cadence. However, the mean stance and swing phases are significantly different for the right side, but not for the left. PD patients have predominantly larger swing phases on the right side, and NPH on the left side (Fig.2H).

For the symmetry ratios Max/Min, none of the values display any significant differences (Fig. S1M-P). This indicates that there is no difference in the overall amount of asymmetry between PD and NPH groups, both displaying asymmetry in some patients. Conversely, for the symmetry ratios R/L (Fig. S1I-L), which are sensitive to which side (R or L) displays asymmetry, the symmetry ratios R/L of mean stance phase (Fig. 2G) and mean swing phase (Fig. 2H) are different between the PD and NPH groups with statistical significance. However, this difference lies in the asymmetry being in opposite directions for the two groups, not in one group being more asymmetric than the other (for these parameters). Asymmetry was further measured in an alternative way (Fig. S3), as described extensively in Methods Sec. 4.6. For each gait cycle parameter, the values for the left foot and the right foot were compared with a statistical test to assess significant differences. Both methods to quantify gait asymmetry were then compared by overlapping in Fig. S4 the results of this second approach to the values of the symmetry ratios (Fig. S2), illustrating that they are complementary and yield consistent results.

**Figure 2:**
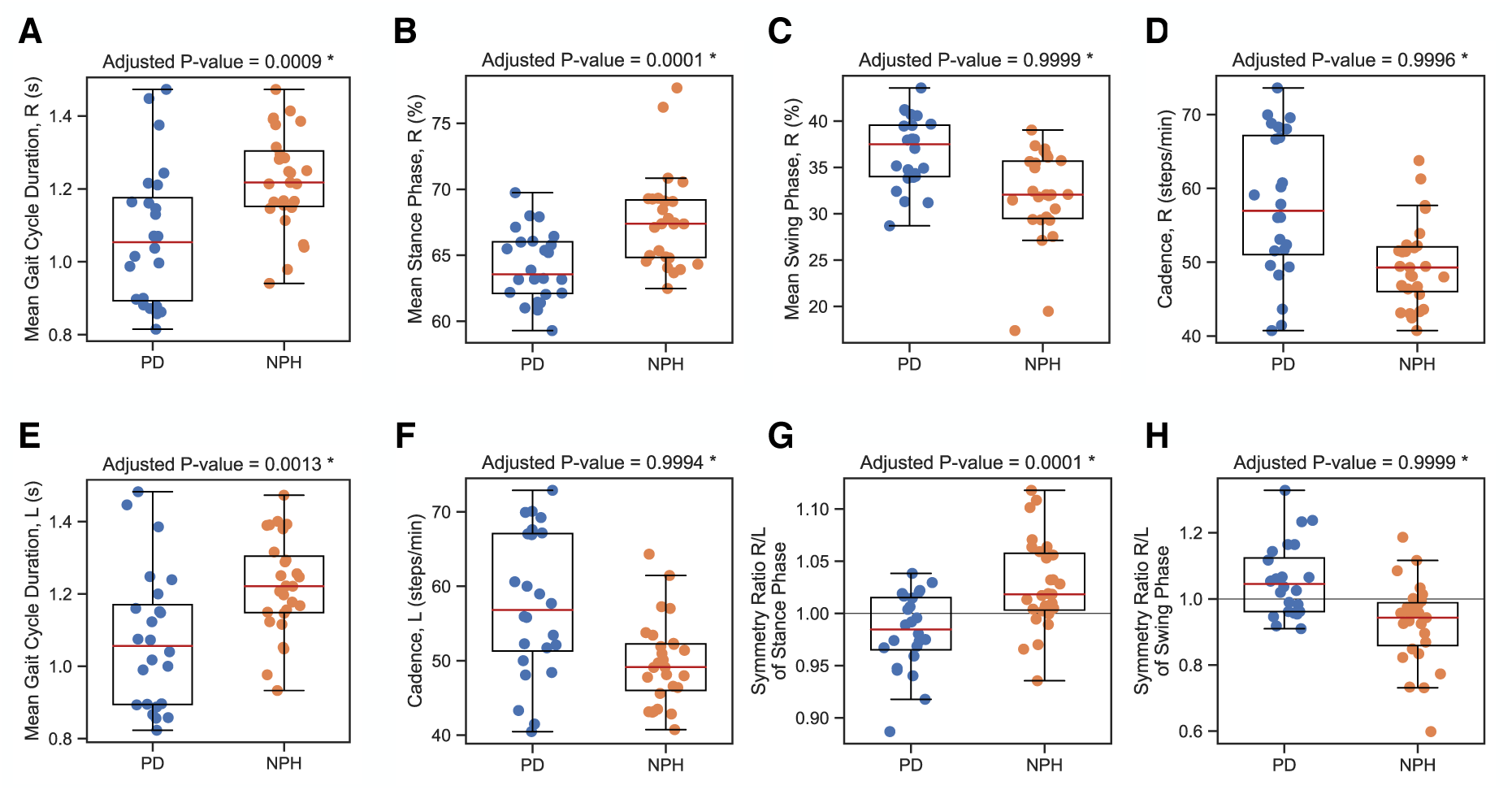
Significantly different features from the statistical comparison of extracted features (gait cycle parameters and symmetry ratios) between PD and NPH patients. Each dot represents the value of the corresponding extracted feature for the gait of one patient performing a ten meters walk. PD patients are displayed in blue (on the left side of each panel), and NPH patients in orange (on the right side). A box plot is superimposed over the data points of each group. **(A)** Mean gait cycle duration right, **(B)** mean stance phase right, **(C)** mean swing phase right, **(D)** cadence right, **(E)** mean gait cycle duration left, **(F)** cadence left, **(G)** symmetry ratio R/L of mean stance phase, **(H)** symmetry ratio R/L of mean swing phase. In all panels, p-values were computed by Welch t-test and corrected for multiple hypothesis testing using Bonferroni correction, see Methods Sec. 4.7. Asterisks indicate that the groups of PD and NPH patients are different with statistical significance (threshold of 0.05) after correction. The complete statistical comparison with all sixteen extracted features is in Supplementary Material Fig. S1. In panels G and H, the horizontal black line indicates the value of the symmetry ratios corresponding to perfect symmetry.

### 2.3 Machine learning classification of PD versus NPH

In this work, several ML models were trained for the classification of PD and NPH patients using the 16 features described above (Table 2). Details can be found in Methods Sec. 4.8. A principal component analysis (PCA) did not fully separate PD and NPH patients (Fig. S5A, B). A uniform manifold approximation and projection (UMAP) obtained similar results (Fig. S5C, D). Hence, we trained ML models for the classification of these two disease groups. Given the small cohort size, data were split into train, validation, and test sets. A nested cross-validation with a leave-pair-out scheme in the outer loop and a leave-one-out scheme in the inner loop was used (Figs. 3A and S6). The inner loop tunes the hyperparameters of the ML model and selects the most relevant features to distinguish the gait between PD and NPH. These are then used by the outer loop. In total, there were 132 models trained in the outer loop, each time tested with a different couple of patients (one PD and one NPH) that was left out from the training and validation.

**Figure 3:**
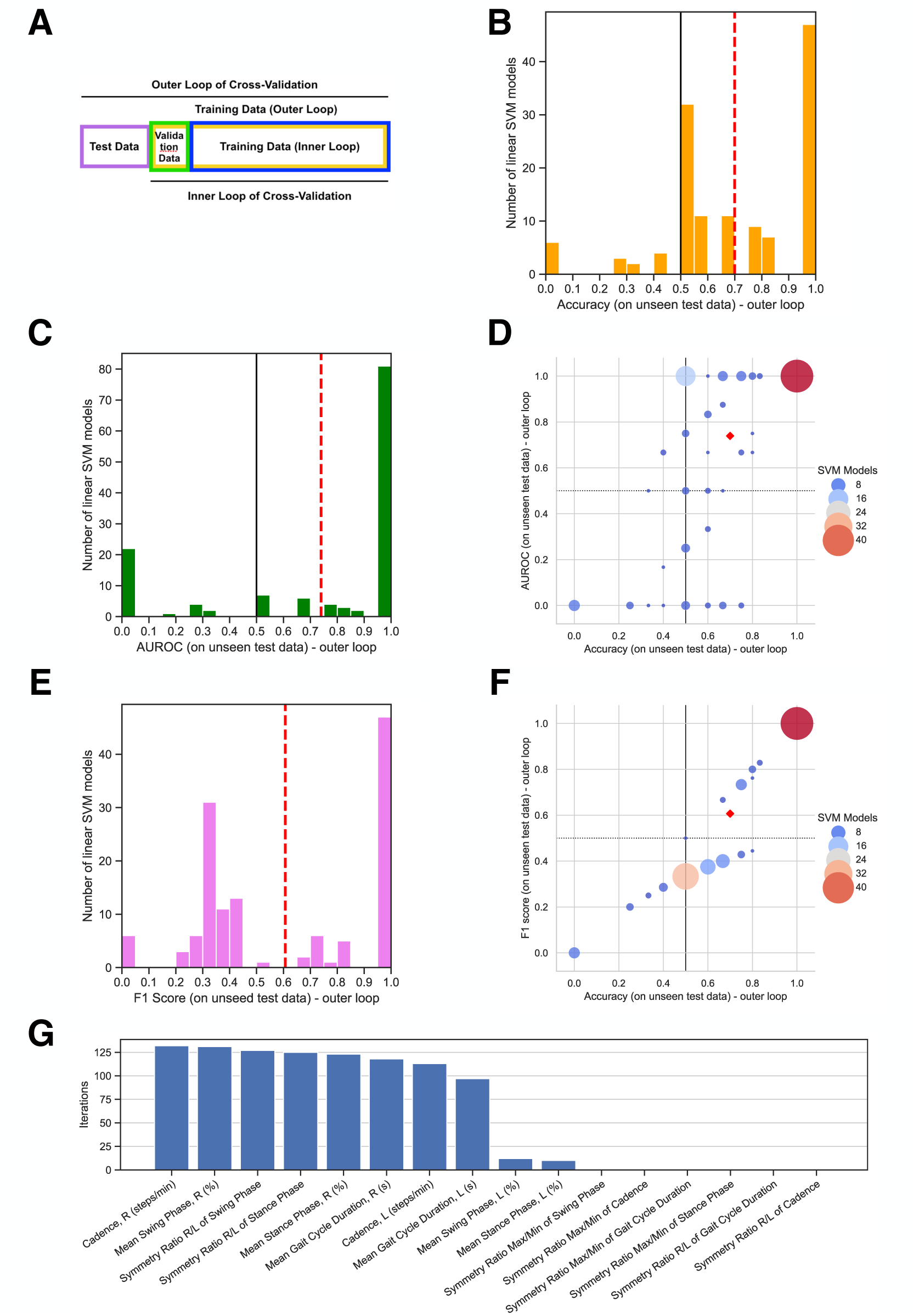
Machine learning (ML) classification of PD versus NPH. **(A)** Data separation into training (20 patients), validation (one patient) and test (one PD and one NPH patients), for the nested cross-validation. **(B)** Distribution of accuracy on unseen test data for the 132 models (SVM with linear kernel) trained in the outer loop of the nested cross-validation. The red line indicates the average value over all models, the black line the value of 0.5 (random classification). **(C)** Distribution of area under the ROC curves (AUROCs). **(D)** Distribution of the 132 models in the plane Accuracy vs AUROC. The red diamond indicates average values. **(E)** Distribution of F1 score on unseen test data. **(F)** Distribution of the 132 models in the plane Accuracy vs F1 score. **(G)** Ranking of all features w.r.t. how often they are retained by the ML algorithm as useful to distinguish between PD and NPH.

Twenty-seven different ML classifiers were tested. Support vector machines with linear kernels (SVM) resulted among the top performers according to several metrics (Fig. S7) and therefore were chosen as the main classification tool. Linear SVM also leads to interpretable models, a reason which further supports this choice. The distribution over the 132 models of the classification accuracy on the unseen test data in the outer loop of the nested cross-validation is shown in Fig. 3B, and has an average of 0.70 and standard deviation of 0.28. The distribution over the 132 models of the area under the ROC curve (AUROC) on the unseen test data can be seen in Fig. 3C. The average AUROC on the test data is 0.74 with a standard deviation of 0.38. More details are provided in Fig. S8. The distribution of the average classification accuracy on the validation data in the inner loop can be seen in Fig. S9. The large standard deviations indicate that these results changed considerably when a different splitting of the patients into the various data subsets was performed, which is due to a high patient-to-patient variability and to the small number of patients. Still, the high accuracy and AUROC mean values showcase the potential to distinguish PD from NPH patients using ML.

We display how the models are distributed in the plane representing Accuracy versus AUROC in Fig. 3D. The distribution over the models of the F1 score can be seen in Fig. 3E. The average F1 score on the test data is 0.61 with a standard deviation of 0.33. We display how the models are distributed in the plane representing Accuracy versus F1 score in Fig. 3F.

Finally, Fig. 3G displays the ranking of the extracted features in terms of how much they are useful in classifying PD versus NPH with SVM. This is quantified by the number of SVM models (vertical axis) that retained each feature in the feature selection step of the training. Out of the total of sixteen features, eight are kept for most of the models, while the other eight are rarely or never maintained. The eight most retained are: cadence R and L, mean swing phase R, symmetry ratio R/L of swing phase, symmetry ratio R/L of stance phase, mean stance phase R, and mean gait cycle duration R and L. These are exactly the same features for which a statistically significant difference between the distribution of PD and NPH data were observed in the statistical comparison of Fig. 2 in Sec. 2.2. These results underline the consistency and robustness of the identification of the most relevant features for PD versus NPH classification between the statistical comparison and the ML approach.

## 3 Discussion

### 3.1 Overview and interpretation of results

This feasibility study developed a methodology to distinguish PD from NPH using prototype wearable sensors and ML. The wearable sensors consisted of a three-axis gyroscope, a three-axis accelerometer, and eight pressure sensors embedded in each patient’s shoe. Wearable sensors represent a cost-efficient and portable alternative to instrumented treadmills, so their use can be foreseen in clinician’s offices or home monitoring. The implemented pipeline extracted eight gait cycle parameters [33] and eight symmetry ratios [34], for a total of sixteen features. The wearables and pipeline were calibrated and validated using healthy subjects.

The gait pattern in NPH is distinguishable from that of PD, as found in [35]. That latter study used an instrumented treadmill provided with force sensors, which detects spatial disposition of the feet. In contrast, this work does not contain this information since it uses unobtrusive sensors on patients’ shoes, which are not able to identify the relative position between the two feet and their orientation. Still, these sensors can extract gait features. Out of the sixteen features computed, eight had statistically significant differencens between the groups of PD and NPH patients. Moreover, for some patients in each of the two disease groups, gait asymmetry was identified. PD is known to induce gait asymmetry [36], while this is much less clear for NPH.

Support vector machines with linear kernel obtained a classification accuracy of 0.70 ± 0.28, an area under the ROC curve of 0.74 ± 0.39 and an F1 score of 0.61 ± 0.33 at distinguishing PD from NPH patients. The mean values indicate that there is potential to distinguish PD from NPH by using gait patterns. Large standard deviations indicate high dependency of the results on which patients’ gait data are used for algorithm training, mainly due to the limited cohort size. The use of nested cross-validation helps to ensure the robustness and reliability of this result, despite the small size of the cohort, which causes the large standard deviation observed.

### 3.2 Limitations

This study has some limitations. The main one is the small number of patients in the cohort. Collecting more data for training should improve the accuracy and especially the standard deviation of the ML models. The current results are promising, and a larger cohort can lead to more reliable and generalizable models. Another limitation was that the wearable sensors, being embedded in the shoes of the subjects, only measured temporal parameters (e.g. gait cycle duration), not spatial ones (like step length). This is a disadvantage compared to equipped treadmills in the identification of NPH, since NPH patients are known to exhibit outwardly rotated feet and abnormal distance between the steps. On the other hand, wearable sensors are cost-efficient and can be used in either clinical or home settings. Finally, no patient with additional diseases was in the cohort, e.g. patients with orthopedic problems, spinal diseases or vascular problems. So, within this study, it is not possible to conclude if for those groups the differentiation would be possible, and how accurate this could be.

### 3.3 Outlook

A larger number of patients is likely to lead to higher performance algorithms. These can then be used at home by taking advantage of the wearable nature of the sensors. This aligns with the general trend to reduce hospital stays and costs, and enhance remote patient monitoring [37, 38, 39, 40, 41, 42]. This is especially relevant in rural areas or countries with a high square milage, where patients might live hours away from their clinician. The pipeline could also be used to quantify disease progression, by repeating the walking movements periodically. Finally, it is likely that adding more information such as using other wearable sensors, genetic and imaging data, will improve the performance of the ML algorithms.

## 4 Methods

### 4.1 Study design and population

This work consists of three complementary studies, which together demonstrate the feasibility of distinguishing gait impairment differences between PD and NPH patients by using wearable sensors and a computational pipeline for gait cycle parameters extraction. The first two studies validate the ensemble of sensors and computational pipeline for gait cycle parameters extraction, and the third one is a clinical study that applies the developed approach on PD and NPH patients in a clinical setting. The first study aims to validate foot pressure sensors against an instrumented treadmill with integrated force sensors, assessing how precisely the ActiSense device measures classical gait parameters. The second study analyzes and evaluates how asymmetry affects the gait of healthy subjects and determines if asymmetry can be reliably measured with the sensors and pipeline used in this work. The third study aims to distinguish gait between the two disease groups (building upon knowledge from the first two studies). Ethical approval for data collection and written informed consent was obtained from all participants in each of the three studies. Lack of major orthopedic problems was a condition for healthy subjects and clinical patients to be included in the study. Each subject wore standardized shoes. Demographic and clinical characteristics of both study groups are shown in Table 1. The three studies are as follows:

**System validation study (SVS).** The goal of SVS was to calibrate and validate the ensemble of sensors and computational pipeline on healthy subjects against an instrumented treadmill, which represents the gold standard in the measurement of gait cycle parameters. Nine healthy subjects were recruited among students at Trier University of Applied Sciences, Trier, Germany. Their characteristics are reported in the second column of Table 1. Each subject was equipped with wearable sensors and performed three one minute-long walks on the instrumented treadmill, each walk at a different speed, respectively of 2 km/h, 3 km/h and 4 km/h. The Gaitway 3D instrumented treadmill (h/p cosmos sports & medical GmbH, Bavaria, Germany), located in the department of physiotherapy at the Trier University of Applied Sciences, was used. The main findings of SVS are reported in Sec. 4.4, Fig. S10, S11 and S12, and an additional detailed description is provided in [43], including exact measurement protocols.

**Asymmetry validation study (AVS).** The goal of AVS was to assess the potential of the ensemble of sensors plus computational pipeline to detect gait asymmetry. Twelve healthy subjects were recruited among graduate students and employees at the University of Luxembourg, Belval, Luxembourg. Their characteristics are reported in the third column of Table 1. Walking tests: each participant was asked to repeatedly perform a clinically standardized gait test, consisting of a 10 m walk. Three tasks were performed by each patient: 10 m walk with a mass of 2.6 kg attached to the left calf by velcro straps, 10 m walk with the same mass attached to the right calf, and 10 m walk without weights. Subjects walked at a self-paced speed, denoting the preferred comfortable gait speed of the individual. Each task was repeated twice, for a total of six 10 m walks for each patient. The weights were used to induce gait asymmetry, similarly to what was done in [44]. The main findings of AVS are reported in Sec. 4.5, Fig. S13 and S14, and an additional detailed description of AVS is provided in [43], including exact measurement protocols.

**Clinical study (CS).** The goal of CS was to assess the potential of the system to distinguish PD from NPH patients by their gait cycle impairments in a clinical setting. Subjects measured: 12 PD and 11 NPH patients were recruited at the National Department of Neurosurgery at the Centre Hospitalier de Luxembourg (CHL), Luxembourg City, Luxembourg. Their characteristics are reported in the fourth and fifth columns of Table 1. Details of individual PD and NPH patients are summarized in Table S1 and S2. Each participant was asked to repeatedly perform a clinically standardized gait test, consisting of a 10 m walk, at their maximum possible walking speed (without running). The task was repeated twice, for a total of two 10 m walks for each patient. The findings of CS are reported in Fig. 2, 3, S1, S3, S4, S2, S5, S6, S7, S8, and S9.

### 4.2 Wearable sensors

For all studies in this work, the ActiSense device (IEE S.A., Luxembourg [45], Fig. 4A) was used to measure gait. The non-invasive foot sensor prototype consists of a sensor housing with an electronic control unit (ECU, Fig. 4A), to which a shoe insole is connected to measure foot pressure by means of 8-high-dynamic pressure cells (Fig. 4B). An inertial measurement unit (IMU, Fig. 4A) containing a three-axis accelerometer, a three-axis gyroscope and a three-aies magnetometer is also embedded in the ECU. The ActiSense system’s IMU signals are internally synchronized with the foot pressure signals on the ECU. Measurements were started and stopped via Bluetooth using IEE’s Smartphone app. A range of shoes (sizes from 37 to 45) of the same brand were purchased and the insoles inserted. Each subject was then given a suitable pair of shoes and the ECU was clipped to the outside of the shoe. For subsequent analysis, only the signals from the pressure sensors, the accelerometer and the gyroscope are employed. For each subject, there are 28 measurement channels given by acceleration in the direction of the 3-axis (Fig. 4C), angular velocity around the 3-axis (Fig. 4D), and pressure from the 8 pressure cells, multiplied by 2 (left and right foot). Example data are displayed for selected channels for the acceleration in Fig. 4E, for the angular velocity around the z-axis in Fig. 4F, and for the pressure in Fig. 4G. Foot pressure and IMU sensor data were recorded at a frequency of 200 Hz, read out from the internal sensor memory (16 GB) and analyzed offline with custom written software, primarily using Python (Version 3.8.8) and also MATLAB™(R2023b, MathWorks Inc., USA).

**Figure 4:**
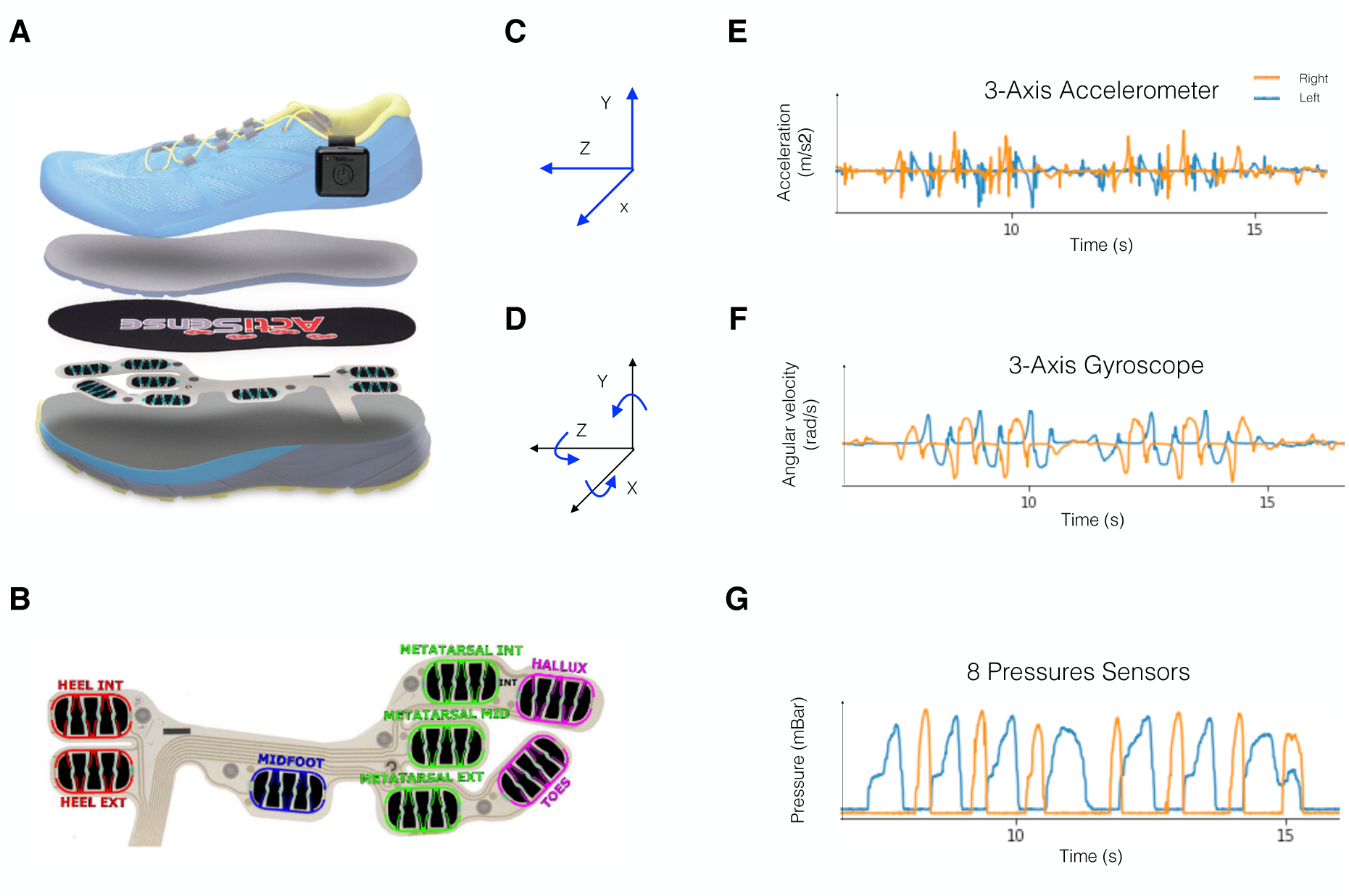
Wearable sensors used in this study: IEE’s ActiSense system [45]. **(A)** Inertial measurement unit (IMU) and pressure sensors located in the shoe. **(B)** Insole with eight pressure sensors [45]. **(C)** The accelerometer measures acceleration along three-axis. **(D)** The gyroscope measures angular velocities around the three-axis. **(E)** Example of accelerometer time-series data. **(F)** Example of gyroscope time-series data. **(G)** Example of pressure time-series data for one sample channel.

### 4.3 Computational pipeline for gait cycle parameter extraction

A computational python pipeline was developed to extract gait cycle parameters, largely implementing and adapting algorithms from [33]. The time-series raw data recorded over 28 channels by the ActiSense system are first filtered by a low-pass filter (IMU data) or Gaussian filter (Pressure data). Then, “toe on” (TON*_i_*) and “heel off” (HOFF*_i_*) events are identified from the gyroscope z-axis signal, for each step *i*. Next, “heel on” (HON*_i_*) and “toe off” (TOFF*_i_*) events are identified from the pressure data. For these four types of events, we use the same definitions as [33]. The events HON*_i_*, HOFF*_i_*, TON*_i_* and TOFF*_i_* are expressed in physical units of time (and measured in seconds).

From these four types of events, the computational pipeline developed computes subsequently the gait cycle parameters in the time domain, namely average gait cycle duration (GCD), average stance phase (STP), average swing phase (SWP) and cadence (C), for left (*L*) and for right (*R*) foot. The definitions of these quantities, clarifying how they are computed, are reported in Table 3. An additional index taking the values of L or R is used as in e.g. HON*_k,i_* to respectively indicate Right (*k* = *R*) or left (*k* = *L*). The index *i* indicates the step for which the quantity is calculated, so *i* + 1 is the subsequent step with respect to *i*. For any of the gait cycle parameters above, mean values are calculated as 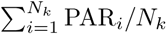, where *N_k_* is the number of steps taken on side *k*, PAR*_i_* is any of GCD*_k,i_*, STP*_k,i_*, SWP*_k,i_*, C*_k,i_* and *k* is either *R* or *L*. Mean stance phase and mean swing phase are expressed as percentages of the corresponding GCD. The same definitions as in [43] were used.

**Table 3:** Definitions of extracted features. Lines 1-8: gait cycle parameters, defined as in [33]. Lines 9-16: symmetry ratios of the gait cycle parameters, defined as in [34]. These definitions are further explained in Methods Sec. 4.3 and discussed in more detail in [43]. A summary of this table can be found in Table 2. Abbreviations are as follows: right (*R*), left (*L*), “heel on” (HON), “heel off” (HOFF), “toe on” (TON), “toe off” (TOFF).

| Extracted Feature | Definition |
| --- | --- |
| Mean gait cycle duration, right ($\text{GCD}_R$) | $\sum_{i=1}^{N_R} (\text{HON}_{R,i+1} - \text{HON}_{R,i}) / N_R$ |
| Mean stance phase, right ($\text{STP}_R$) | $\sum_{i=1}^{N_R} (\text{TOFF}_{R,i} - \text{HON}_{R,i}) / N_R \cdot 100 / \text{GCD}_R$ |
| Mean swing phase, right ($\text{SWP}_R$) | $\sum_{i=1}^{N_R} (\text{HON}_{R,i+1} - \text{TOFF}_{R,i}) / N_R \cdot 100 / \text{GCD}_R$ |
| Cadence, right ($\text{C}_R$) | $N_R / (\text{HON}_{R,N_R+1} - \text{HON}_{R,1})$ |
| Mean gait cycle duration, left ($\text{GCD}_L$) | $\sum_{i=1}^{N_L} (\text{HON}_{L,i+1} - \text{HON}_{L,i}) / N_L$ |
| Mean stance phase, left ($\text{STP}_L$) | $\sum_{i=1}^{N_L} (\text{TOFF}_{L,i} - \text{HON}_{L,i}) / N_L \cdot 100 / \text{GCD}_L$ |
| Mean swing phase, left ($\text{SWP}_L$) | $\sum_{i=1}^{N_L} (\text{HON}_{L,i+1} - \text{TOFF}_{L,i}) / N_L \cdot 100 / \text{GCD}_L$ |
| Cadence, left ($\text{C}_L$) | $N_L / (\text{HON}_{L,N_L+1} - \text{HON}_{L,1})$ |
| Symmetry ratio R/L of GCD | $\text{GCD}_R / \text{GCD}_L$ |
| Symmetry ratio R/L of STP | $\text{STP}_R / \text{STP}_L$ |
| Symmetry ratio R/L of SWP | $\text{SWP}_R / \text{SWP}_L$ |
| Symmetry ratio R/L of C | $\text{C}_R / \text{C}_L$ |
| Symmetry ratio Max/Min of GCD | $\max(\text{GCD}_R, \text{GCD}_L) / \min(\text{GCD}_R, \text{GCD}_L)$ |
| Symmetry ratio Max/Min of STP | $\max(\text{STP}_R, \text{STP}_L) / \min(\text{STP}_R, \text{STP}_L)$ |
| Symmetry ratio Max/Min of SWP | $\max(\text{SWP}_R, \text{SWP}_L) / \min(\text{SWP}_R, \text{SWP}_L)$ |
| Symmetry ratio Max/Min of C | $\max(\text{C}_R, \text{C}_L) / \min(\text{C}_R, \text{C}_L)$ |

In addition, for each parameter the corresponding symmetry ratio R/L is computed (by taking the ratio parameter right / parameter left), which was found by [34] to be the best metric to assess gait cycle asymmetry. An alternative version of the symmetry ratio is also computed, namely symmetry ratio Max/Min, where the largest gait cycle parameter value was always kept at the numerator. By definition, this ratio thus assumes only values equal to 1 (symmetry), or larger than 1 (asymmetry), but doesn’t provide information on which side (left or right) has a larger parameter. This makes up a total of 16 gait cycle parameters and symmetry ratios (Table 2), to be used as features for the subsequent statistical analysis and classification by machine learning.

### 4.4 System validation study (SVS): validating sensors and pipeline against treadmill on healthy subjects

A system validation study (SVS) was performed to calibrate and validate the sensors and computational pipeline on nine healthy subjects (Table 1) against an instrumented treadmill. The sensors and pipeline were tested on healthy young controls to have a baseline calibration and validation. Each subject was provided with wearable sensors and performed three, one-minute-long walks on an instrumented treadmill, each walking at a different speed, respectively of 2, 3 and 4 km/h. More details are provided in Methods Sec. 4.1 and in [43]. The results of SVS are shown in Fig. S10, Fig. S11 and Fig. S12. Each panel in Fig. S10 represents a comparison between the measurements performed by the wearable sensors and by the treadmill for one of the four gait cycle parameters computed (A: mean gait cycle duration, B: cadence, C: mean stance phase and D: mean swing phase; definitions in Methods Sec. 4.3 and Table 2). The comparison was repeated for each of the different subjects and different speeds. For each subject and speed, the discrepancy between the measurement performed by the wearable system and by the treadmill was computed. This is reported in Fig. S11, panels A-D (for the four respective gait cycle parameters), where the discrepancy is referred to as *Error* and computed as *Error* ≐ 100 · (*Par_Wearable_* − *Par_Treadmill_*) */Par_Treadmill_* for *Par* indicating any of the four parameters mentioned above. It is reported in Fig. S12, panels A-D, the average *Error* over all subjects for each speed, and the overall average for subjects and speeds (each panel corresponding to a gait cycle parameter).

In Fig. S10A and B, the values of mean gait cycle duration and cadence between the wearable sensors and the treadmill are compared. The discrepancies between these values are quantified in Fig. S11A and B, where the percent error between wearable sensors and treadmill values for mean gait cycle duration and cadence are reported for each subject and speed. Treadmill values (representing the ground truth) have been centered at 0. Overall, for mean gait cycle duration and cadence, the errors in all subjects and speeds are less than 1% for mean gait cycle duration, and less than 2% for cadence. In Fig. S12A, B, the average percent error between wearable sensors and treadmill values for mean gait cycle duration and cadence for every speed is reported, in addition to the average overall speeds. The average errors are less than 0.2% for mean gait cycle duration and less than 0.3% for cadence. In Fig. S10A and B, values of mean gait cycle duration for both systems decrease roughly linearly with increasing speed, while cadence values increase roughly linearly, as expected. In Fig. S10A, the standard deviation (quantifying the spread of gait cycle values across different gait cycles for each subject and speed) is only slightly larger for the wearables with respect to than that of the treadmill, indicating that the precision by which the wearable system measures these two quantities is only slightly lower than that of the treadmill. Overall, for mean gait cycle duration and cadence, there is a close agreement between wearable sensors and the treadmill. It can be concluded that the wearable sensors and treadmill measure these quantities similarly well for all practical purposes.

For both values of mean stance phase and mean swing phase, there was initially a systematic discrepancy between the values obtained from the treadmill and those from the wearable sensors. This shift was in the same direction and of roughly the same magnitude for each subject and speed (and different for mean swing and stance phases). This was expected and the source of this systematic shift is identified in the filtering of the raw time-series data for the pressure sensors. It was corrected by introducing in the pipeline an a posteriori calibration of the mean stance and swing phase values. This is thoroughly described in [43]. In Fig. S10C and D, the comparison between measurements from the wearable sensors and treadmill for mean stance and swing phase after calibration is displayed. After calibration, the values of the system are close to the treadmill values. The discrepancy between these values is quantified in Fig. S11C and D. The percentage error between wearable sensors and treadmill values for mean stance phase and mean swing phase are reported for each subject and speed. Overall, for mean stance phase and mean swing phase, an agreement between wearable sensors and treadmill values is found, with the errors in all cases (all subjects and speeds) being less than 6% for mean stance phase, and less than 15% for mean swing phase. Fig. S12C, D report the average percentage error between wearable sensors and treadmill values for mean stance phase, mean swing phase for every speed, and the average over all speeds. These average errors are less than 3% for mean stance phases and less than 5% for mean swing phase. In Fig. S10C, D, for each subject, values of mean stance phase and mean swing phase for both systems decrease roughly linearly with increasing speed, as is expected. In Fig. S10C, D, the standard deviation (quantifying the spread of stance and swing phase values across different gait cycles for each subject and speed) is only slightly larger for the wearable sensors with respect to the treadmill, indicating that the precision by which the wearable system measures these two quantities is only slightly lower than that of the treadmill. Overall, for mean stance and swing phases (after calibration) an agreement can be seen between the wearable sensors and the treadmill.

### 4.5 Asymmetry validation study (AVS): assessing system’s capability to measure gait asymmetry in healthy subjects

The asymmetry validation study (AVS) was performed to assess the potential of the sensors and pipeline to detect gait asymmetry. This was performed on 12 healthy subjects (Table 1). Each participant was asked to perform two repetitions for each of the following three tasks: 10 m walk with a mass of 2.6 kg attached to the left calf (red datapoints in Fig. S13 and Fig. S14), 10 m walk with a mass of 2.6 kg attached to the right calf (blue datapoints), and 10 m walk without weights (green data points), see [43] for an extended description. The weights were used to induce gait asymmetry, similarly to what was done in [44].

In Fig. S13 and Fig. S14 the results for the symmetry ratios of the four gait cycle parameters (symmetry ratio of gait cycle duration, cadence, stance phase, and swing phase) are displayed. Each panel reports the symmetry ratio for the first repetition of the 10 m walk on the x-axis, and the value for the second repetition on the y-axis. A value of 1 indicates perfect symmetry, smaller/larger than 1 indicates that the gait cycle parameter investigated is larger for the left/right foot. The two repetitions have the role of technical replicates, so it was expected that the two repetitions would yield similar results for all cases, and that all data points were to be located roughly along the diagonal. In both figures, this is the case for most data points. This represents a first consistency check of the capability for the system to yield consistent, reproducible measurements of gait asymmetry across repeated trials.

In Fig. S14, the symmetry ratios of gait cycle duration (A) and cadence (B) for all tasks (weights left, weights right, no weight) and all subjects are very close to the value of 1. This is expected as cadence and gait cycle duration measured on the left or right foot should be very similar in the conditions of this test. Cadence counts the steps on one side, and gait cycle duration measures the time between one heel strike and the successive heel strike from the same foot. This represents a further consistency check for the system that, while attempting to measure asymmetry, does not measure noticeable asymmetry in quantities which are not supposed to display any.

In Fig. S13, the symmetry ratios for stance phase (A) and swing phase (B) are displayed. In both cases, the three groups (weights left, red, weights right, blue, no weight, green) are well spread apart, almost disjointed from each other (the overlap being only for very few subjects). The subjects are around the value of (1,1) when walking with no weight (green), and further away when walking with weights left (red) or right (blue). The shift of each group with weights occurs in opposite directions between panel A (stance phase) and panel B (swing phase). This is because the stance phase (foot touching the ground) and the swing phase (foot not touching the ground) are complementary, in that their sum must be equal to the gait cycle duration. If the stance phase increases on one side due to added weight, the corresponding swing phase decreases. This is evidence that the system (sensors plus pipeline) has a very high potential to detect and quantify asymmetry in gait cycle parameters, such as stance phase and swing phase. Swing phase occupies a smaller fraction of the gait cycle than stance phase. The spread of the data points is larger when measuring the symmetry ratio of swing phase (panel B), making swing phase the best parameter to detect gait asymmetry. In panel A, healthy subjects walking with no weight have a symmetry ratio of swing phase roughly within the range 0.9-1.1, while when walking with weights, the symmetry ratio is roughly above 1.1 or below 0.9. It can be concluded that the system displays a very high potential to detect asymmetry in swing phase, distinguishing between physiological asymmetry (0.9-1.1), and pathological asymmetry (below 0.9 or above 1.1).

### 4.6 Gait asymmetry detection in PD and NPH patients

With the developed system, gait asymmetry between PD and NPH was investigated. Key findings are described in Sec. 2.2, while here additional details are provided, and it is stressed how an alternative method to assess gait asymmetry was used as well. This led to results consistent with those from the symmetry ratios, thus further supporting the validity of such an approach.

Gait asymmetry in PD and NPH patients was characterized in Fig. S1I-P, Sec. 4.6 and Fig. S2, where the symmetry ratios are shown for mean gait cycle duration, mean stance phase, mean swing phase, and cadence (computed as in Tab. 2 for the patients of the CS in Tab. 1). Overall, it was found that in both PD and NPH groups, there were patients exhibiting gait asymmetry that the system can detect. For PD, this is not surprising because, while it is known that gait dynamics in healthy young subjects is mostly symmetric [46], PD is known to lead to gait asymmetry [36]. It has also been argued that PD might usually start from the left hemisphere [47], however the left- or right-handedness of the patient should be considered. Of particular interest is Fig. S1K, where the symmetry ratio of swing phase is displayed, and based on the calibration (Methods Sec. 4.5), values outside the range 0.9-1.1 indicate pathological asymmetry. Patients with asymmetry in swing phase at a pathological level (i.e. outside the range 0.9-1.1) are present in both PD and NPH groups (Fig. S1K). This asymmetry appears in opposite directions for PD and NPH, and this difference can only be appreciated in the symmetry ratio (R/L), not in the symmetry ratio (Max/Min).

None of the Max/Min ratios display any significant difference (Fig. S1), indicating that there is no difference in the amount of asymmetry between PD and NPH groups. However, in Fig. 2H, both groups contain some walks from some patients which turn out to display pathological gait asymmetry, as they have an asymmetry index on swing phase outside the window of 0.9-1.1. This window was identified in the Asymmetry Validation Study (AVS), Methods Sec. 4.5, to contain values indicating physiological gait asymmetry, while values outside this window indicate pathological gait asymmetry. When considering the symmetry ratios R/L for stance and swing phase (which are sensitive to which side has larger value, unlike the symmetry ratios Max/Min), both symmetry ratios for stance phase R/L and for swing phase R/L are significantly different between PD and NPH groups, with PD having predominantly larger swing phases on the right side, and NPH on the left side. It is worth mentioning here that PD is known to induce gait asymmetry, while this is much less clear for NPH.

The comparison in gait asymmetry between PD and NPH in this study can be further deepened by looking at the significant differences in the gait cycle parameters between right and left Fig. S3. This is an alternative approach to using the symmetry ratio, and Fig. S4 shows their equivalence. To assess the statistical significance of gait asymmetry, asymmetry was further measured in an alternative way. In Fig. S3, the values for mean gait cycle parameters for each patient (of both groups PD and NPH) and each walk are displayed. The mean values (across all steps performed during each walk) and the standard deviations (measuring how spread the values were between the different steps of a walk) are considered. A Welch t-test was performed to compare each value of a right gait cycle parameter with the corresponding left value, with Bonferroni correction for multiple hypothesis testing.

The difference between left and right mean gait cycle duration was never significant between PD and NPH (Fig. S3). For PD, 17% of all walks display a statistically significant difference in mean stance phase between left and right. In 4 walks, the left mean stance phase was significantly larger. Correspondingly, 17% of all walks display a statistically significant difference in mean swing phase between left and right, and in 4 walks the right mean swing phase was significantly larger. For NPH, 26% of all walks display a statistically significant difference in mean stance phase between left and right. In 6 walks, the right mean stance phase was significantly larger, and only in 1 patient was it larger on the left side. Correspondingly, 26% of all walks display a statistically significant difference in mean swing phase between left and right. In 6 walks, the left mean swing phase was significantly larger, and only in 1 patient was it larger on the right side.

Both methods to quantify gait asymmetry were compared by overlapping the data (Fig. S4), illustrating that they are complementary and yield consistent results. The walks displaying significant left/right asymmetry appear in the tails of the distributions of symmetry ratios (Fig. S4C-F). This supports the validity of the symmetry ratio used here to quantify gait asymmetry. Particularly in Fig. S4E and F, walks that have a statistically significant difference between left and right swing phase (according to the analysis of Fig. S3E and F) lead to a symmetry ratio that is smaller than 0.9 or larger than 1.1 (as indicated by the calibration of SVS, Sec. 4.5), further supporting the validity of the range identified as pathological.

In Sec. 4.5 asymmetry (on swing phase symmetry ratio) is within 0.9-1.1 for young healthy subjects and when artificially impaired with weights (or when considering PD and NPH), values fall instead mostly outside this range. This shows that the system (sensors plus pipeline) can measure both physiological and pathological gait asymmetry and distinguish between the two. (to some extent). This cohort of patients is too small to draw conclusions on the disease. However, it is an important result that shows that with this approach, asymmetry of gait is detectable in these types of patients. This will help to further the investigation of this subject in the future.

### 4.7 Statistical data analysis

Statistical significance of differences between the means of extracted features (for any of the sixteen features of Table 2) is assessed by Welch t-test. The tests are performed two-tailed. This is used to compare extracted features between PD and NPH patients in Fig. 2 and Fig. S1 and to compare right and left values in Fig. S3A and B. In all cases, p-values are corrected for multiple hypothesis testing by Bonferroni correction. Corrected p-values are compared to a threshold of 0.05 to declare statistical significance. For any of the gait cycle parameters in Table 2, as detailed in Table 3 mean values are STP*_k,i_*, SWP*_k,i_*, C*_k,i_* and *k* is either *R* or *L*. Errorbars displayed in Fig. S10A-D, S3A and B represent standard deviations computed as 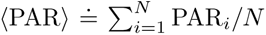, where *N* is the number of steps taken, PAR*_i_* is any of GCD*_k,i_*, STP*_k,i_*, SWP*_k,i_*, C*_k,i_* and *k* is either *R* or *L*. Errorbars displayed in Fig. S10A-D, S3A and B represent standard deviations computed as 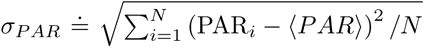. Data visualization in Fig. S5 is performed by principal component analysis (PCA) and uniform manifold approximation & projection (UMAP).

### 4.8 Machine learning

Machine learning (ML) algorithms were used to perform binary classification to distinguish PD from NPH patients data. Eleven PD and twelve NPH patients were considered, Table 1.

#### Extracted Features

For each 10 m walk, the computational pipeline described above computes eight gait cycle parameters, namely gait cycle duration, stance phase, swing phase and cadence, for left and for right foot, as in [33]. In addition, eight symmetry ratios are computed. The symmetry ratio Right/Left for each gait cycle parameter was computed as in [34]. Moreover, an alternative version of the symmetry ratio, namely symmetry ratio Max/Min, also from [34], was further computed, where the largest gait cycle parameter value was always kept at the numerator. Thus, by construction this ratio can only assume values equal to 1 (symmetry), or larger than 1 (asymmetry). It also doesn’t carry the information on which side (left or right) has a larger parameter. This makes a total of 16 extracted features, listed in Table 2. Thus, each 10 m walk led to a data point in a 16 dimensional space (visualized by PCA and UMAP in Fig. S5), in which the ML classifier is run. All features were standardized beforehand by removing their mean and scaling them to unit variance (Z-score).

#### Data separation for cross-validation

When training a ML classifier, a common good practice is to split the data in training and test set, so that the classifier is trained on a part of the data, and its performances are assessed on another part of the data, not used during training. In order to reduce the dependency of this process by the actual realization of this random splitting of the data into training and test set, usually a k-fold cross-validation procedure is also used, where this split and training process is repeated k times, and the performance values are averaged over all models trained. Here, in order to additionally identify the best values for the hyperparameters of each model, a nested cross-validation approach was performed, i.e. the data were split in training set, test set and validation set, Fig. 3A. The nested cross-validation included two loops.

The outer loop uses leave-pair-out cross-validation, splitting the data into unseen test data and training data for the outer loop, and leading to test each model’s performance on the unseen test data. In this leave-pair-out cross-validation loop performed (Fig. S6C, a part of patients (one PD and one NPH, Fig. S6B) is left out as test data, and all repetitions of walks from the same patient are kept in the same partition of the data set). This leads to 11*12 = 132 models being trained in the outer loop, see e.g. Fig. S8C.

Moreover, the training set for the outer loop is further split for each of these models into a training and validation set, using a leave-one-out cross-validation (Fig. S6F) that leaves one patient in turn in the validation set. The goal of this inner loop is to tune the hyperparameters of each model. This inner loop is performed before the outer loop, as the hyperparameters values determined here are then used in the models of the outer loop.

#### Comparison of machine learning classifiers

A total of 27 ML classifiers was run, including support vector machine classifiers (SVM) with linear kernel (linear SVM), on the classification task of PD versus NPH on our data split as above. The performances of such classifiers is compared in terms of how well they perform the classification of PD vs NPH, Fig. S7. The performance of the classification is quantified by several metrics: accuracy (panel A), balanced accuracy (B), F1 score (C) and area under the receiver-operator-characteristic curve (AUROC, D). The SVM with linear kernel is among the top few performers by any of these metrics, with e.g. a balanced accuracy of 0.71. Classifiers are also compared by training time, and SVM also has training time which is shorter or similar to the ones of the other faster methods, Fig. S6E.

#### Support vector machine with linear kernel

For the final PD versus NPH classification, support vector machine classifiers with linear kernel (linear SVMs) were chosen, because of their interpretability, simplicity and performances among the best in the above described comparison. The standard implementation of linear SVM from the python *scikit-learn* package was used. Linear SVM is a well known machine learning model which attempts to linearly classify training data. It has an hyperparameter, C, which weights the relative importance of the number of the misclassified instances and the margins width of the hyperplane. Higher C value means that the number of misclassified samples is low, which can lead to overfitting. A great advantage of linear SVM is that it provides a unique solution [21].

#### Linear SVM hyperparameter tuning, features ranking and performance assessment

In the linear support vector machine classifier (linear SVM), a step is used to rank the features by their importance (Fig. S6D) and to fine-tune the hyperparameter C (Fig. S6E), and the best performing value of C is used for the corresponding SVM model of the outer, leave-pair-out loop (Fig.S6C). The features ranking is performed by random forest ranking the features by the mean decrease in Gini impurity, following the same procedure as [21], and for each iteration of the loop only the top ranked features who have a mean decrease in Gini impurity above the average (over all features) are retained. For the final feature classification of Fig. 3D, for each feature the number of times a feature is retained in any of the 132 models trained in the outer loop is counted, the features are ranked accordingly, with 132 being the highest possible value corresponding to a feature being retained in all iterations, and 0 indicating a feature that is never retained. Thus final feature ranking is obtained by count of occurrences of each feature in the features selection step. In the final classification of Fig. S8C, performance is assessed by computing average accuracy (from the confusion matrix), summarized in Fig. 3B, average area under the ROC curve (AUROC), summarized in Fig. 3C, and average F1 score, summarized in Fig. 3E. The F1 score is computed using the average=’macro’ argument for the F1 score function of the sklearn python package, which simply computes the mean of the F1 score for the PD class and the F1 score for the NPH class.

## Data and codes availability statement

Raw sensor data were collected at the Centre Hospitalier de Luxembourg (CHL). The data for this manuscript is not publicly accessible as it is subject to CHL and its internal regulations. For reasons of patient confidentiality and privacy, anonymised summary data that support the findings of this study are available from the corresponding author or the first authors upon reasonable request and with permission. All computational codes for this project and preprocessed anonymised data necessary to reproduce all results will be made publicly available upon publication on the GitHub repository: https://github.com/StefanoMagni/MoveSenseAI.git.

## Acknowledgements

The clinical study and research work in this manuscript was conducted at the Centre Hospitalier du Luxembourg, to whom we express our gratitude for the support received. The authors would further like to give special thanks to all the study participants. Additionally, we are very grateful to the Luxembourg National Research Fund and the company IEE S.A. for the funding of the MoveSenseAI project that have enabled us to carry out this clinical research study. The authors express their gratitude to Martin Thinnes (IEE S.A., Luxembourg) and Tobias Meyer (IEE S.A.) for the provision and training on the ActiSense device and for sharing their expertise in the field of sensor technology. The authors also thank Prof. Dr. Steffen Müller (Trier University of Applied Sciences, Trier, Germany) for providing us access to the equipped treadmill utilized in this work and for guidance in the design of the corresponding study. Gratitude is also expressed to Prof. Dr.-Ing. Klaus Peter Koch (Trier University of Applied Sciences, Trier, Germany) for enabling the involvement of Konstantinos Verros, who completed his master thesis within the MoveSenseAI project, under the guidance of Dr. Stefano Magni and Dr. Rene Peter Bremm. Moreover, the authors thank Dr. med. Isabel Fernandes Arroteia (Centre Hospitalier du Luxembourg, Luxembourg) for support with clinical measurements and for sharing her clinical expertise in the field of movement disorders. The authors would like to express their deepest gratitude to Dr. Michela Bernini for illustrating the project overview scheme (Fig. 1). The authors are very grateful to Dominique Jeanson for the very valuable assistance in editing and proofreading this manuscript.

## Funding

The research work in the MoveSenseAI project was supported by the Luxembourg National Research Fund (Fonds National de la Recherche, FNR BRIDGES 2020/BM/14772888) and the IEE S.A. company in Luxembourg. Note that all funding bodies played no role in the study design, data collection, analysis and interpretation of data, or the writing of this manuscript.

## Author contributions

Conceptualisation and project coordination: RPB, FH.

Design, study direction, and methodology: SM, RPB.

Organization of clinical study: RPB, FH.

Organization of validation study: KV, RPB, SM.

Sensor-based data collection: RPB, KV, SL.

Data analysis: SM, RPB, KV, XH.

Machine learning: SM, XH, RPB, AH.

Fundings aquisition: RPB, FH, AH.

Supervision: FH, RPB, SM, JG.

Medical interpretation: FH, FJ.

Outreach: SM, KV, RPB.

Writing – Original manuscript: SM, JG, RPB.

Writing – Critical review and manuscript revision: SM, JG, RPB, FH, KV, AH.

All authors have read and agreed to this version of the manuscript.

## Ethics statement

The clinical study was conducted at the Centre Hospitalier de Luxembourg according to the principles of the Declaration of Helsinki (2013). All study participants signed a written informed consent, which included the collection of sensor data. This collection has been approved by the Ministry of Health and National Ethics Board (Comité National d’Ethique de Recherche, CNER, Reference number: 202101/02, 0622-2) in Luxembourg. The validation study of the ActiSense device was carried out on healthy subjects at the Trier University of Applied Sciences according to the principles of the Declaration of Helsinki (2013) with a prior approval of the ethics committee of the State Chamber of Medicine in Rhineland-Palatinate, Germany (Reference name: VitalMove Study).

## Conflict of interests

The authors of this article declare that they have no conflict of interest. The company IEE S.A. contributed to the funding of the MoveSenseAI project in accordance with the terms and conditions of the BRIDGES programme of the Luxembourg National Research Fund.

---

**Figure S1:**
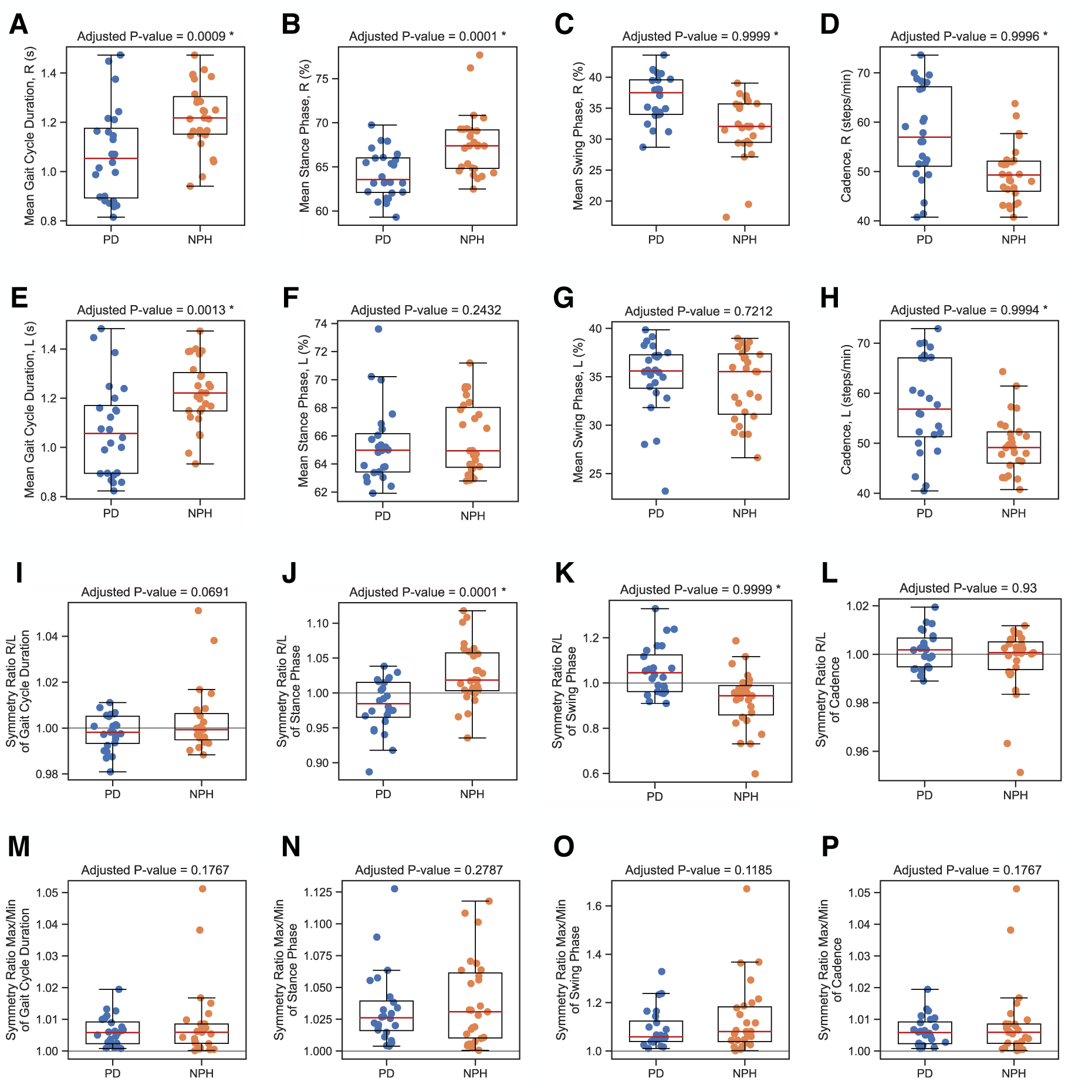
Statistical comparison of extracted features (gait cycle parameters and symmetry ratios) between PD and NPH patients, all features displayed. Each dot represents the value of the corresponding extracted feature for a walk of one patient. PD patients are displayed in blue (on the left side of each panel), and NPH patients in orange (on the right side). A box plot is superimposed over the data points of each group. **(A-D) Mean parameters right. (A)** Mean gait cycle duration right, **(B)** mean stance phase right, **(C)** mean swing phase right, **(D)** cadence right. **(E-H) Mean gait cycle parameters left. (E)** Mean gait cycle duration left, **(F)** mean stance phase left, **(G)** mean swing phase left, **(H)** cadence left. **(I-L) Symmetry ratios R/L, (I)** for mean gait cycle duration, **(J)** for mean stance phase, **(K)** for mean swing phase, **(L)** for cadence. **(M-P) Symmetry ratios Max/Min, (M)** for mean gait cycle duration, **(N)** for mean stance phase, **(O)** for mean swing phase, **(P)** for cadence. In each panel, p-values were computed by Welch t-test and corrected for multiple hypothesis testing by Bonferroni correction, see Methods Sec. 4.7. Asterisks indicates those parameters for which the groups of PD and NPH patients are different with statistical significance (threshold of 0.05, after Bonferroni correction).

**Figure S2:**
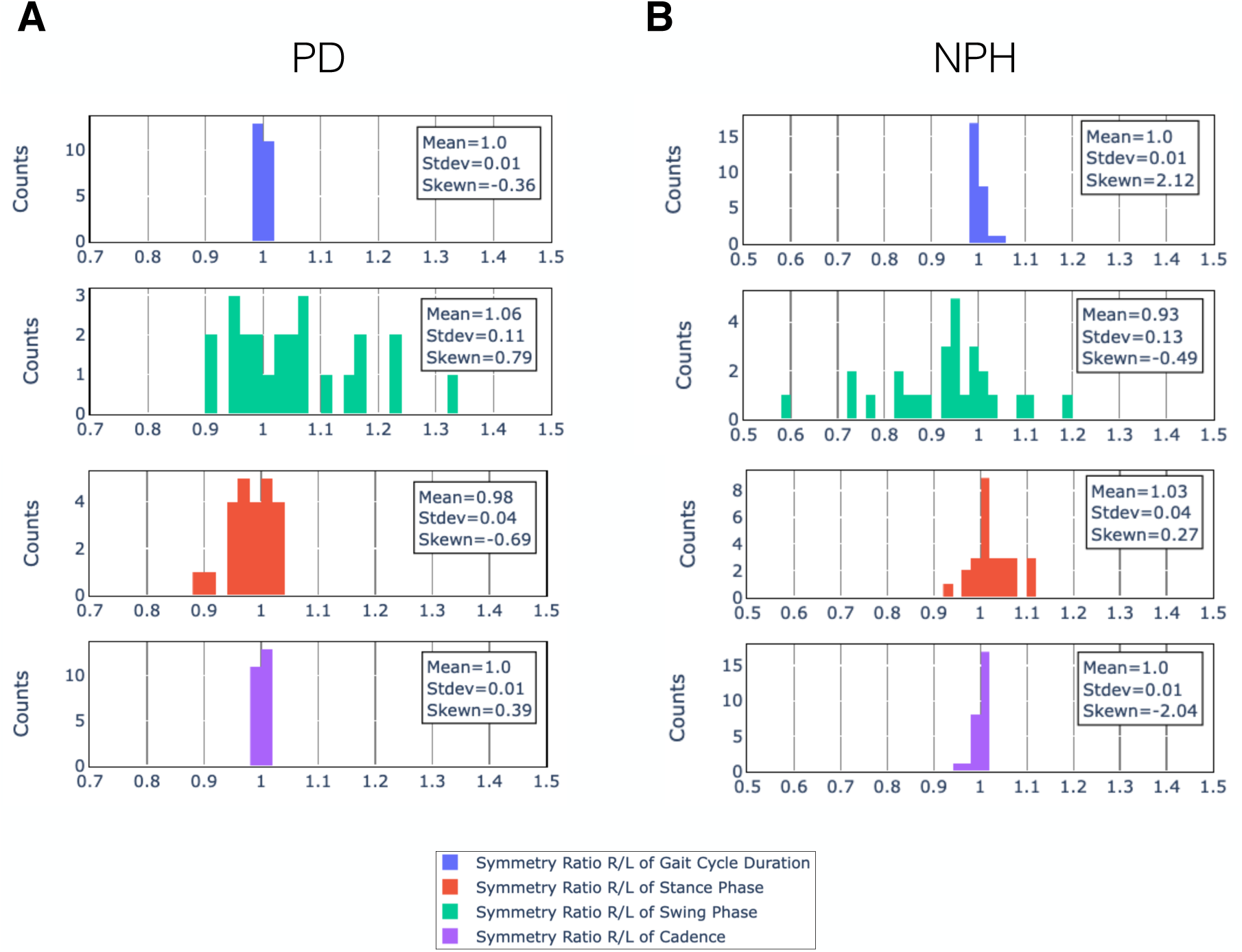
Quantification of gait asymmetry in patients by symmetry ratios R/L, and comparison between PD and NPH. **(A)** Symmetry ratios R/L for PD patients for all gait cycle parameters. **(B)** Symmetry ratios R/L for NPH patients for all gait cycle parameters.

**Figure S3:**
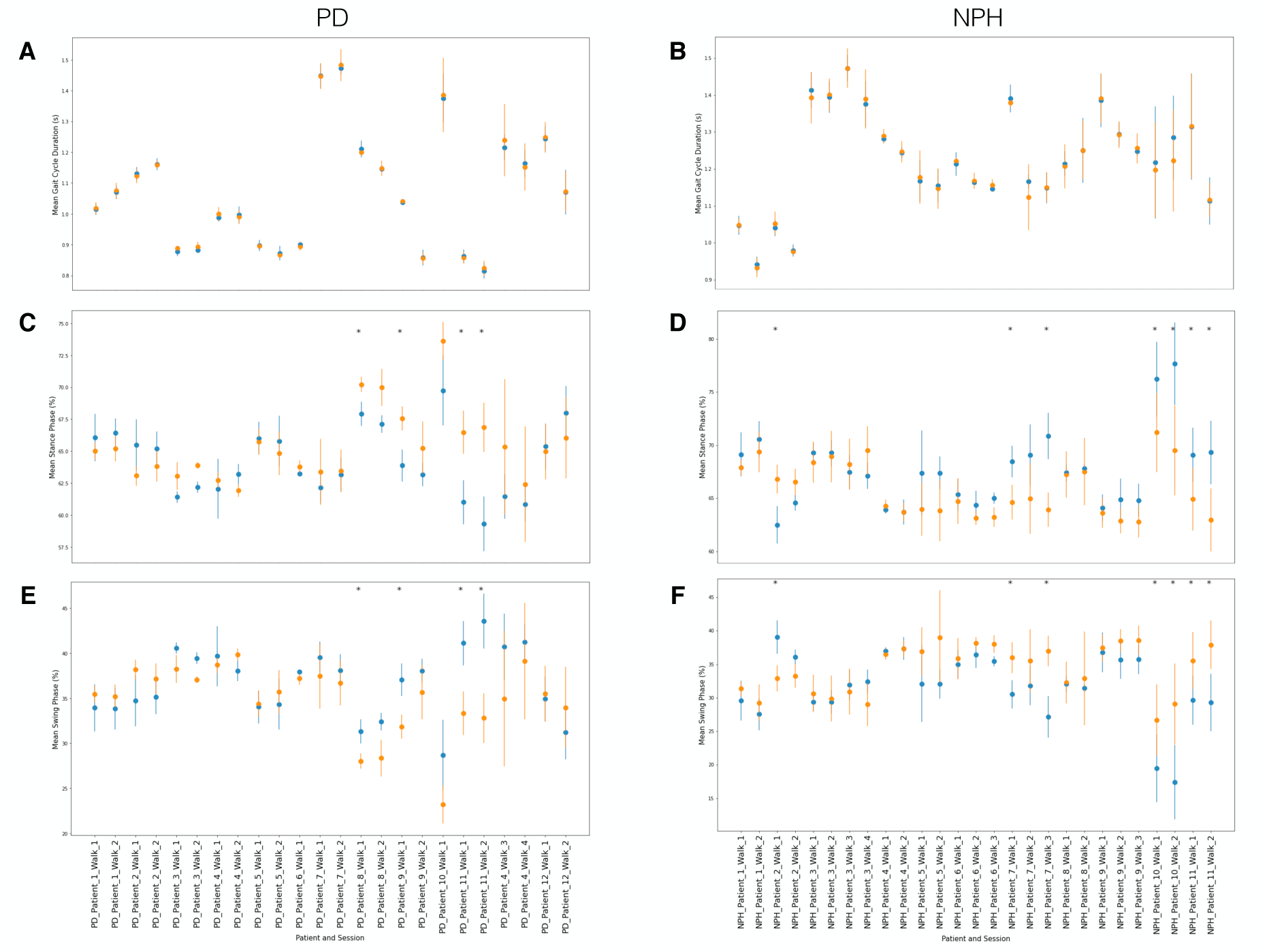
Identification of gait asymmetry in patients by statistical test on gait cycle parameters left (orange dots) vs right (blue dots), and comparison between PD and NPH. **(A)** Comparison between mean gait cycle duration right and left, for PD patients. **(B)** Comparison between mean gait cycle duration right and left, for NPH patients. **(C)** Comparison between mean stance phase right and left, for PD patients. **(D)** Comparison between mean stance phase right and left, for NPH patients. **(E)** Comparison between mean swing phase right and left, for PD patients. **(F)** Comparison between mean swing phase right and left, for NPH patients. In each panel, p-values were computed by Welch t-test and corrected for multiple hypothesis testing by Bonferroni correction, see Methods Sec. 4.7. Asterisks indicates those walks for which the left and right values of a given parameter in a given walk are different with statistical significance (threshold of 0.05, after Bonferroni correction).

**Figure S4:**
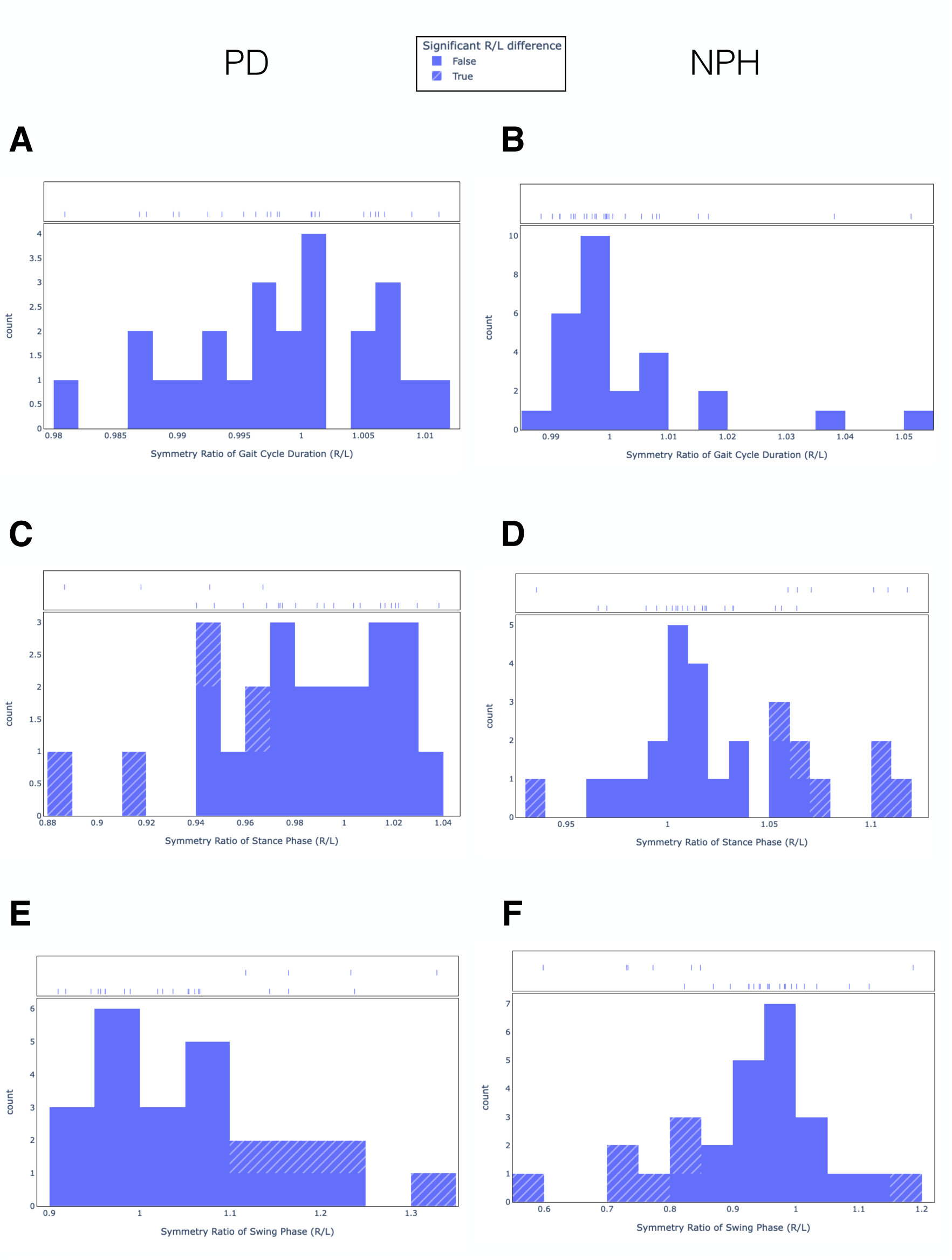
Overlap between significantly different R/L values of gait cycle parameters and corresponding symmetry ratios. **(A)** Overlap of patients with mean gait cycle duration significantly different between R and L and the distribution of symmetry ratio R/L values for gait cycle duration, for PD patients. **(B)** Same, for NPH patients. **(C)** Overlap of patients with mean stance phase significantly different between R and L and the distribution of symmetry ratio R/L values for stance phase, for PD patients. **(D)** Same, for NPH patients. **(E)** Overlap of patients with mean swing phase significantly different between R and L and the distribution of symmetry ratio R/L values for swing phase, for PD patients. **(F)** Same, for NPH patients.

**Figure S5:**
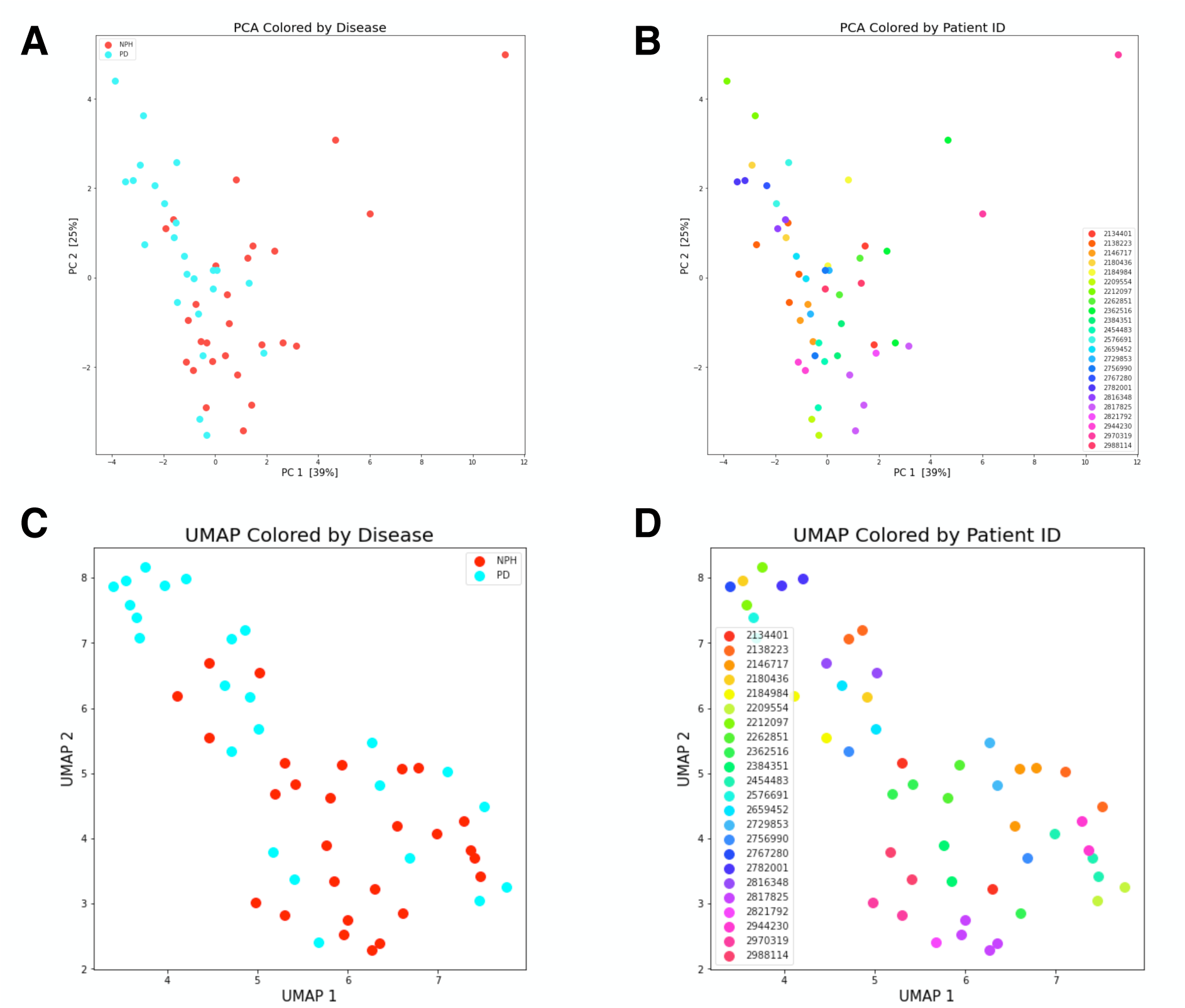
Visualization of PD and NPH data by dimensionality reduction via PCA and UMAP. **(A)** Visualization by Principal Component Analysis (PCA) of the datapoints (sixteen extracted features of Tab. 2) corresponding to PD and NPH patients. **(B)** PCA colored by patient identifier. **(C)** Visualization by uniform manifold approximation & projection (UMAP) corresponding to PD and NPH patients. **(D)** UMAP colored by patient identifier. PCA and UMAP reveal patients’ heterogeneity, with repeated walks from the same patient clustered together (panels B and D). This also showcases the potential (and need) to use systems for individualized follow up of patients in future research.

**Figure S6:**
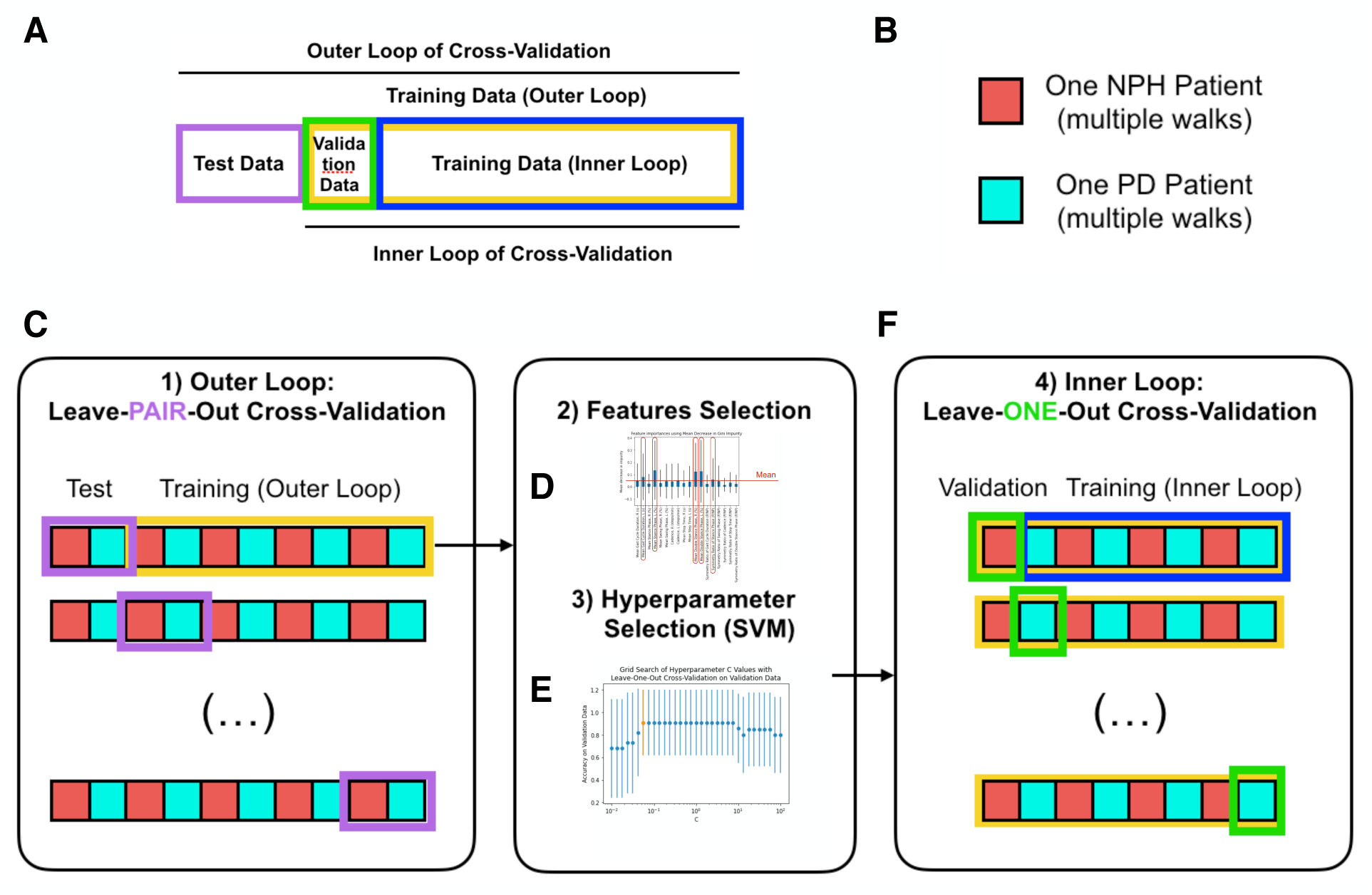
Making the machine learning analysis robust by cross-validation. **(A)** Dataset separation into training, validation and test. **(B)** Data are split by patients, not by walk. **(C)** Leave-pair-out cross-validation of the outer loop. **(D)** Feature selection by Random Forest and mean Gini impurity. **(E)** SVM hyperparameter tuning in the inner cross-validation loop. **(F)** Leave-one-out cross-validation of the inner loop.

**Figure S7:**
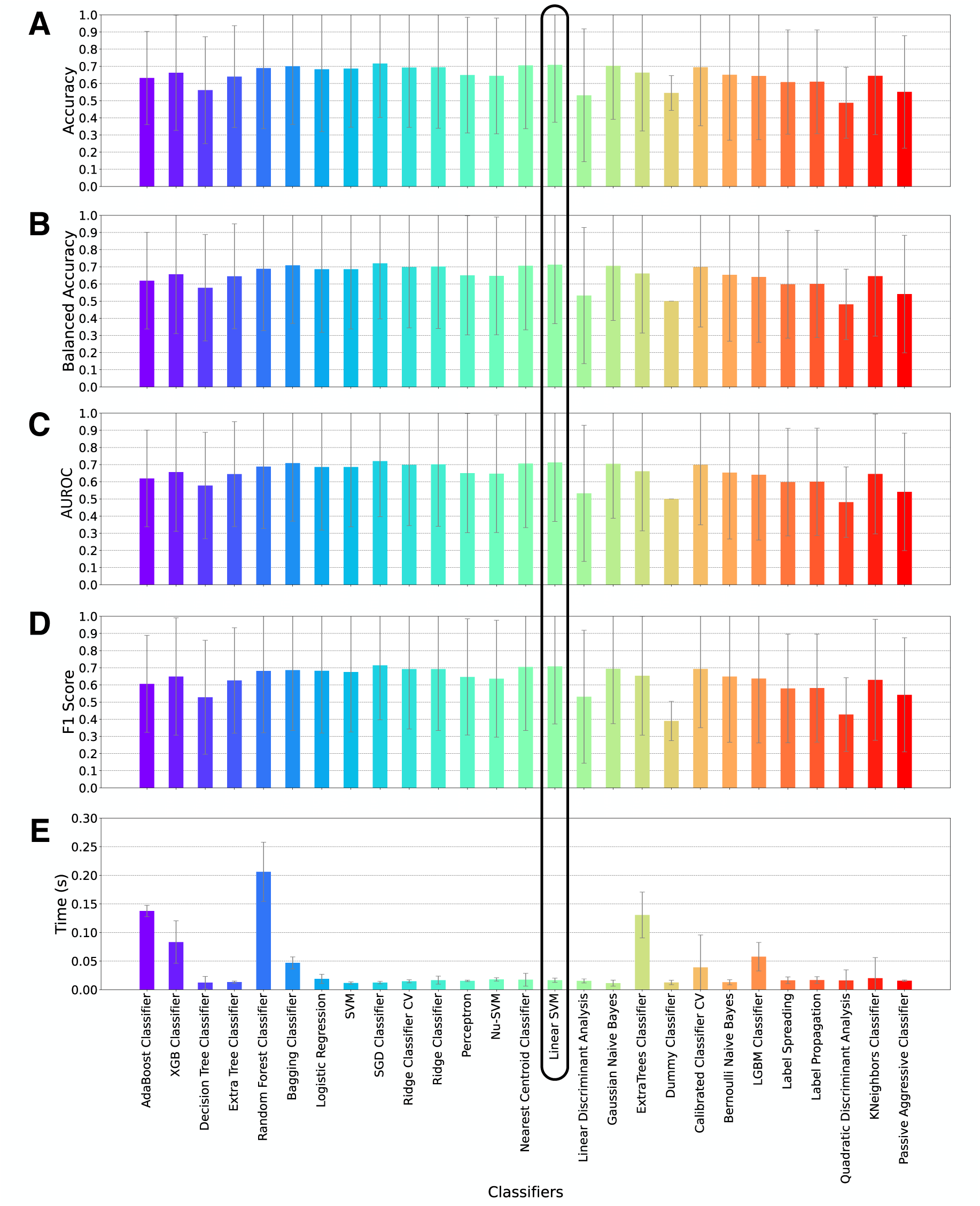
Performance comparison of machine learning algorithms in classifying PD vs NPH patients, all metrics. **(A)** Accuracy. **(B)** Balanced Accuracy. **(C)** Area under the curve of the receiver-operator characteristic (AUROC) curve. **(D)** F1 Score **(E)** Time taken to train each classifier. The black box underlines the classifier used in the clinical study of this paper, namely support vector machine classifier (SVM) with linear kernel.

**Figure S8:**
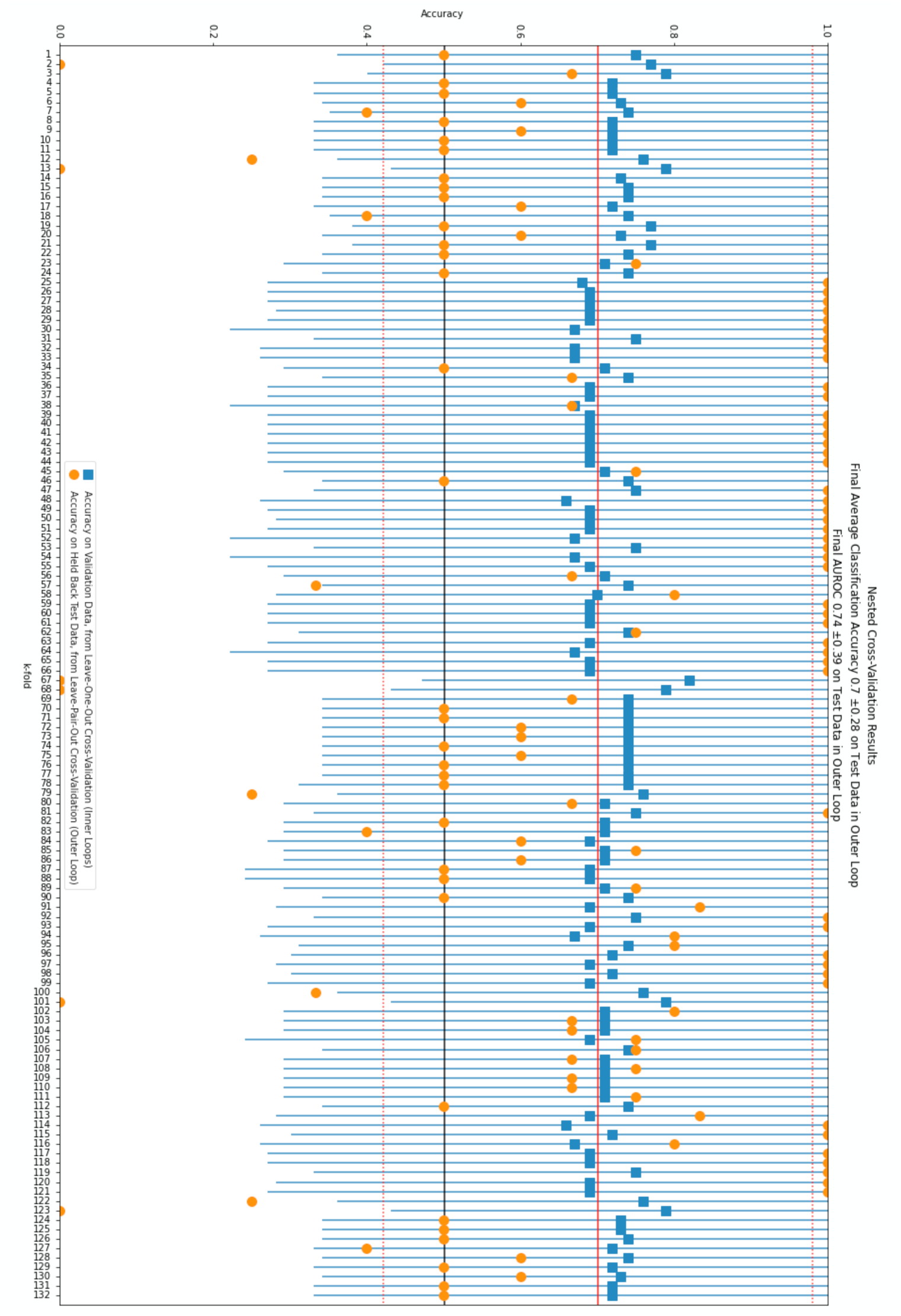
Summary of the performances (accuracies) of all linear support vector machines (SVMs) models trained. A scheme comprising a nested Cross-Validation, with leave-pair-out cross-validation in the outer loop and leave-one-out cross validation in the inner loop was used to train classifiers. For each training in the inner loop, classification accuracies are aggregated and displayed by their mean (blue squares) and standard deviation (blue lines) over all models of each inner loop, which is used to tune the hyperparameter C of the linear SVM classifiers. The outer loop gives rise to 132 linear SVMs models trained (arising from the 12 PD times 11 NPH patients). The 132 models listed on one axis, while the other axis lists accuracies. The accuracy of each of these models is reported as an orange dot. The average of these 132 values is reported as a continuous red line, and the standard deviation as dashed red lines. These 132 values are summarized in Fig. 3B. A solid black line indicates the value of 0.5 for accuracy, corresponding to classifying at random.

**Figure S9:**
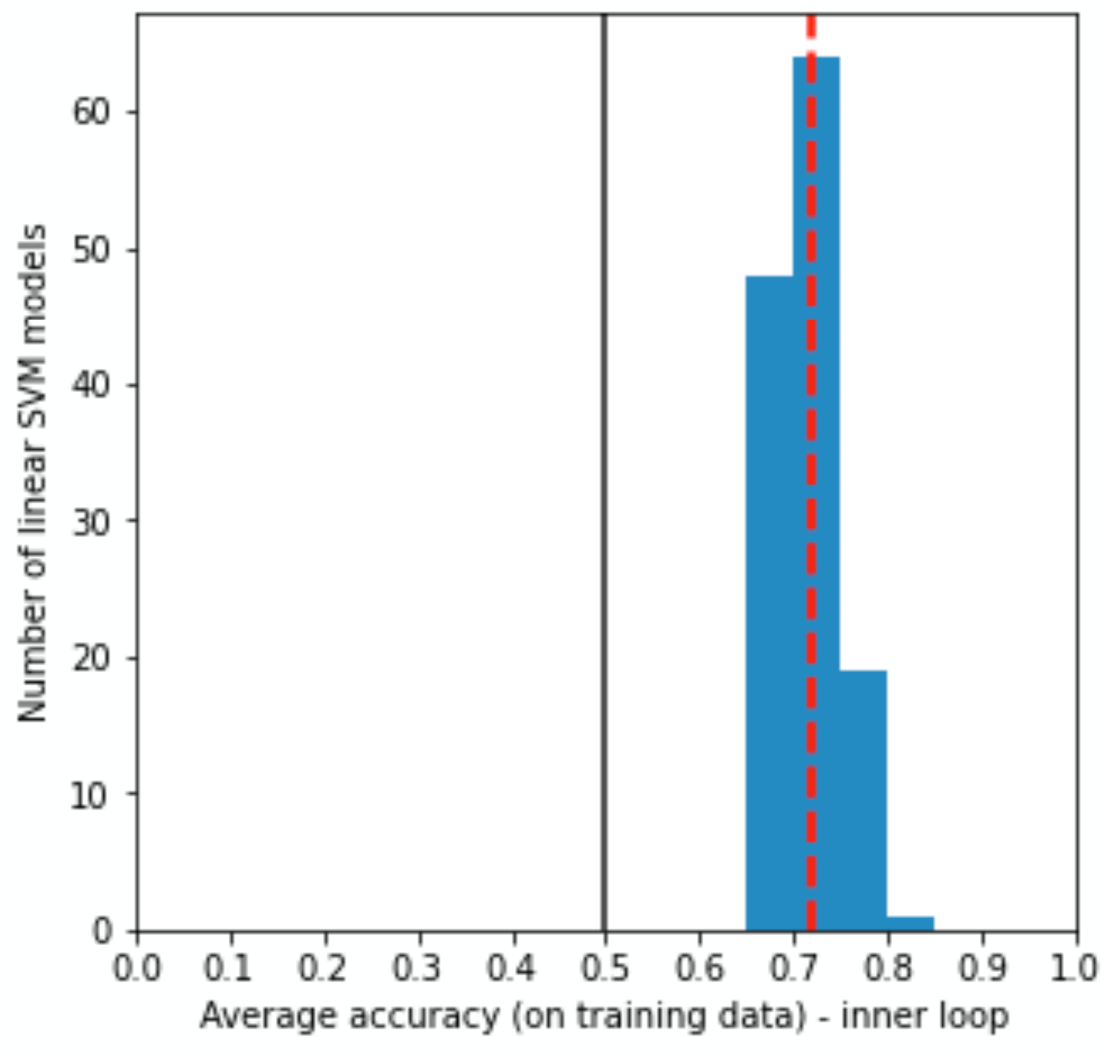
Distribution of average accuracy on training data for the models (SVM with linear kernel) trained in the inner loop of the nested cross-validation scheme. The vertical red line indicates the average value over all models, the vertical black line indicates the value of 0.5 corresponding to a random classification.

**Figure S10:**
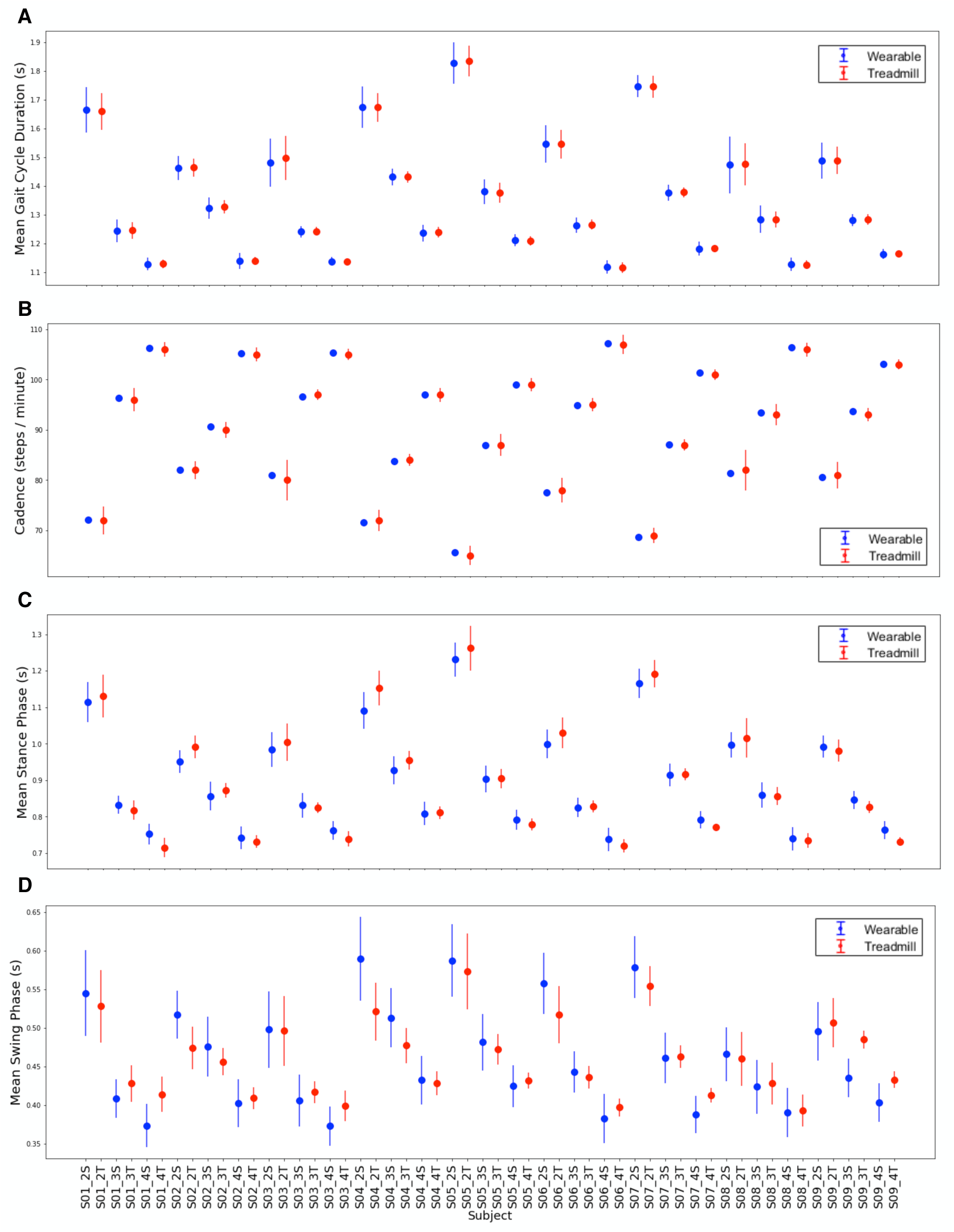
Validation on healthy subjects of the employed system (system validation study, SVS). **(A-D)** System validation with respect to treadmill representing golden standard, see Methods Sec. 4.1. Comparison of wearable with respect to treadmill for **(A)** gait cycle duration, **(B)** cadence, **(C)** stance phase and **(D)** swing phase. SVS is described in Methods Sec. 4.1, and we provided additional details in [43]. Each panel shows for each subject the measurement performed at a given speed by the wearable (blue) and the corresponding measurement performed by the treadmill (red). Each panel has the same order of subjects, and subject labels are SXX-Y-Z, with XX indicating the ID of the subject, Y the speed (in km/h) and Z being either S (for System based on wearables) or T (for Treadmill). Notice that mean stance phase and mean swing phase are defined as times and used as such here however in Methods, Table 3 they are expressed as percentages of the corresponding GCD.

**Figure S11:**
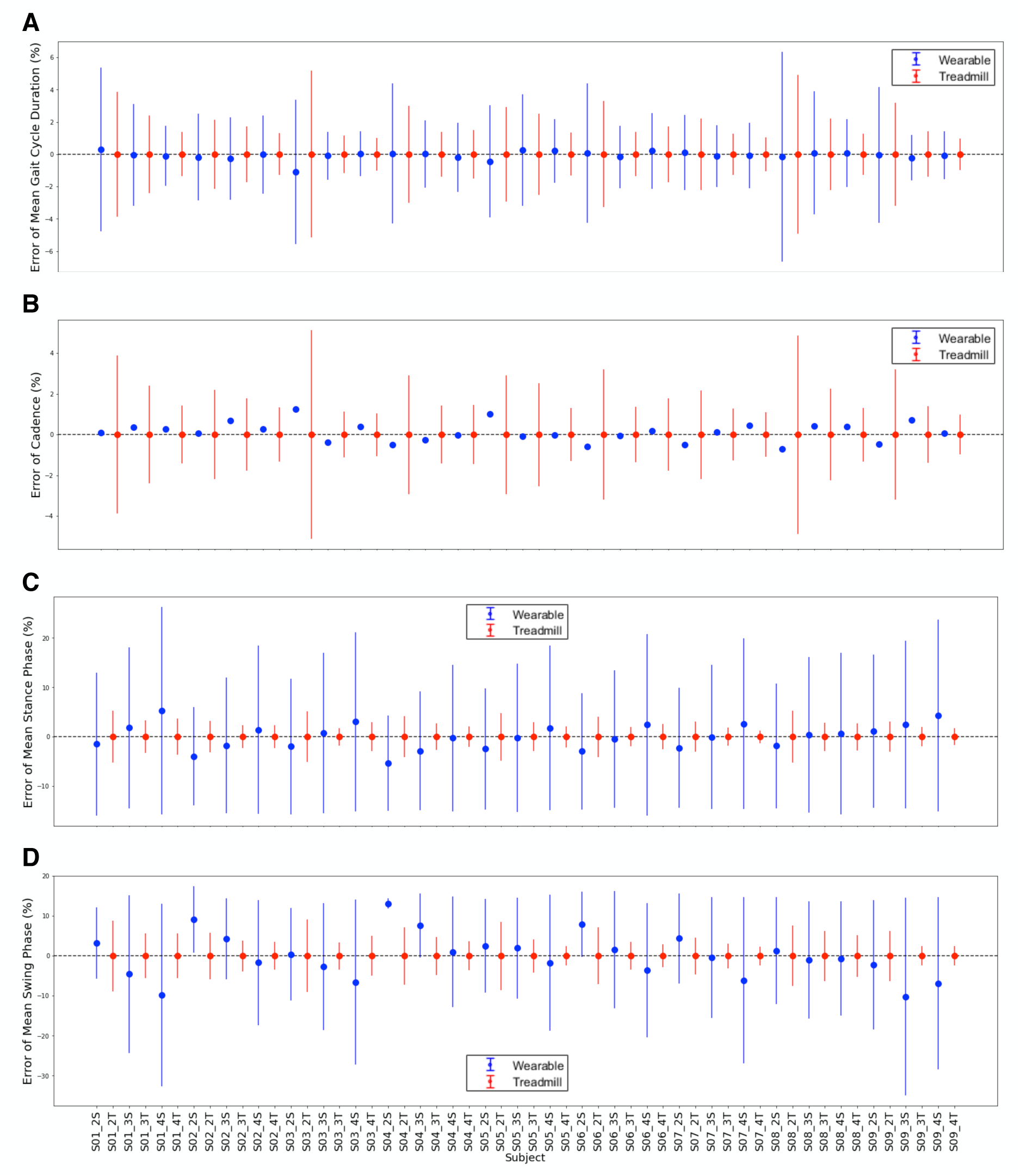
Quantification of differences between measures from wearables and treadmill in the system validation study (SVS), after validation, for each subject. Quantification of error between the measurement performed by our system (wearable plus computational pipeline) and the equipped treadmill, for each subject and for each speed of walking. Errors are displayed for mean gait cycle duration **(A)**, cadence **(B)**, mean stance phase **(C)** and mean swing phase **(D)**.

**Figure S12:**
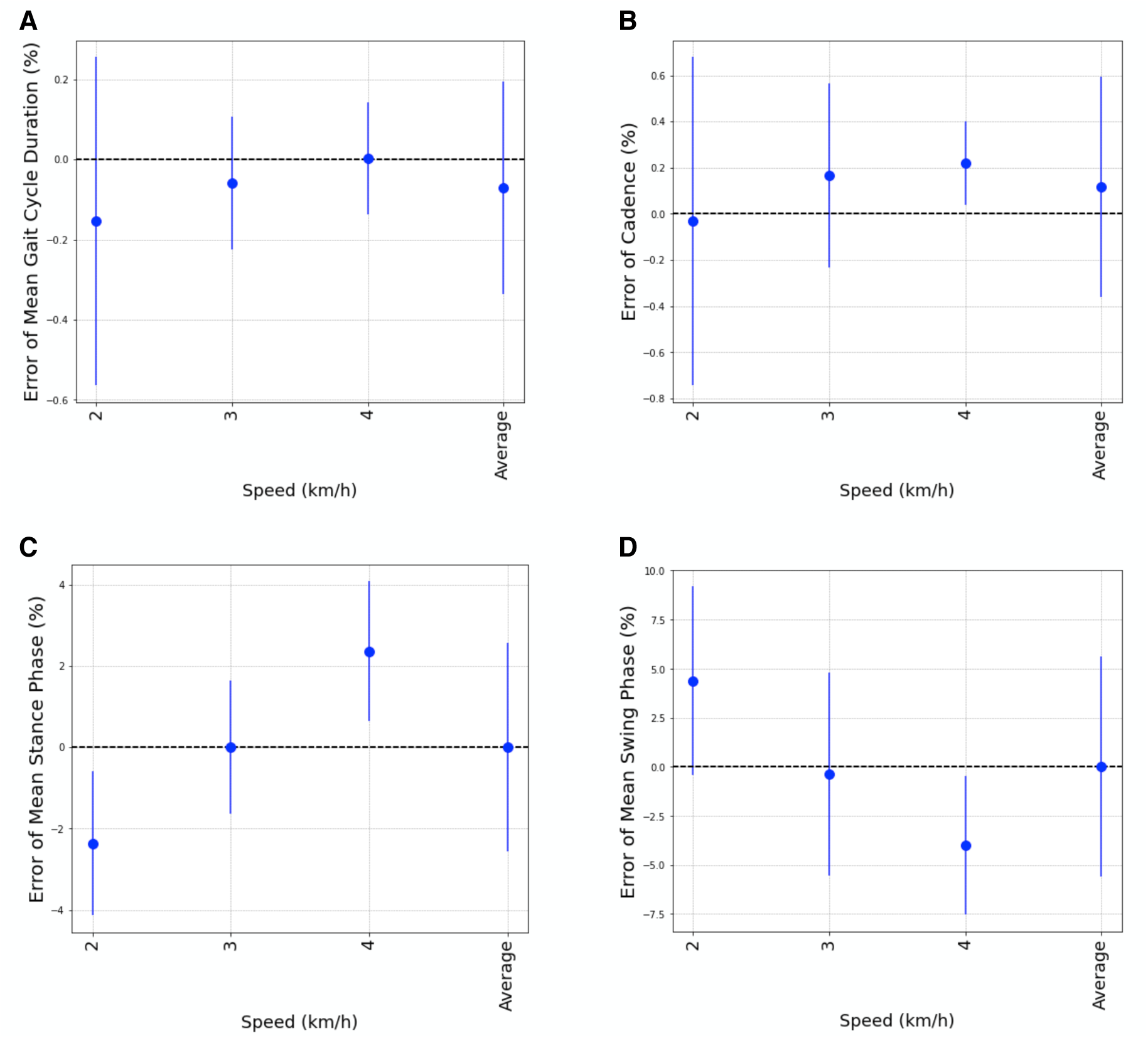
Quantification of differences between measures from wearables and treadmill in the system validation study (SVS), after calibration, averaged over subjects. Quantification of error between the measurement performed by our system (wearable plus computational pipeline) and the equipped treadmill, averaged over subjects and over speeds. Averaged errors are displayed for mean gait cycle duration **(A)**, cadence **(B)**, mean stance phase **(C)** and mean swing phase **(D)**. SVS is described in Fig. S10 and Methods Sec. 4.1, and we provided additional details in [43].

**Figure S13:**
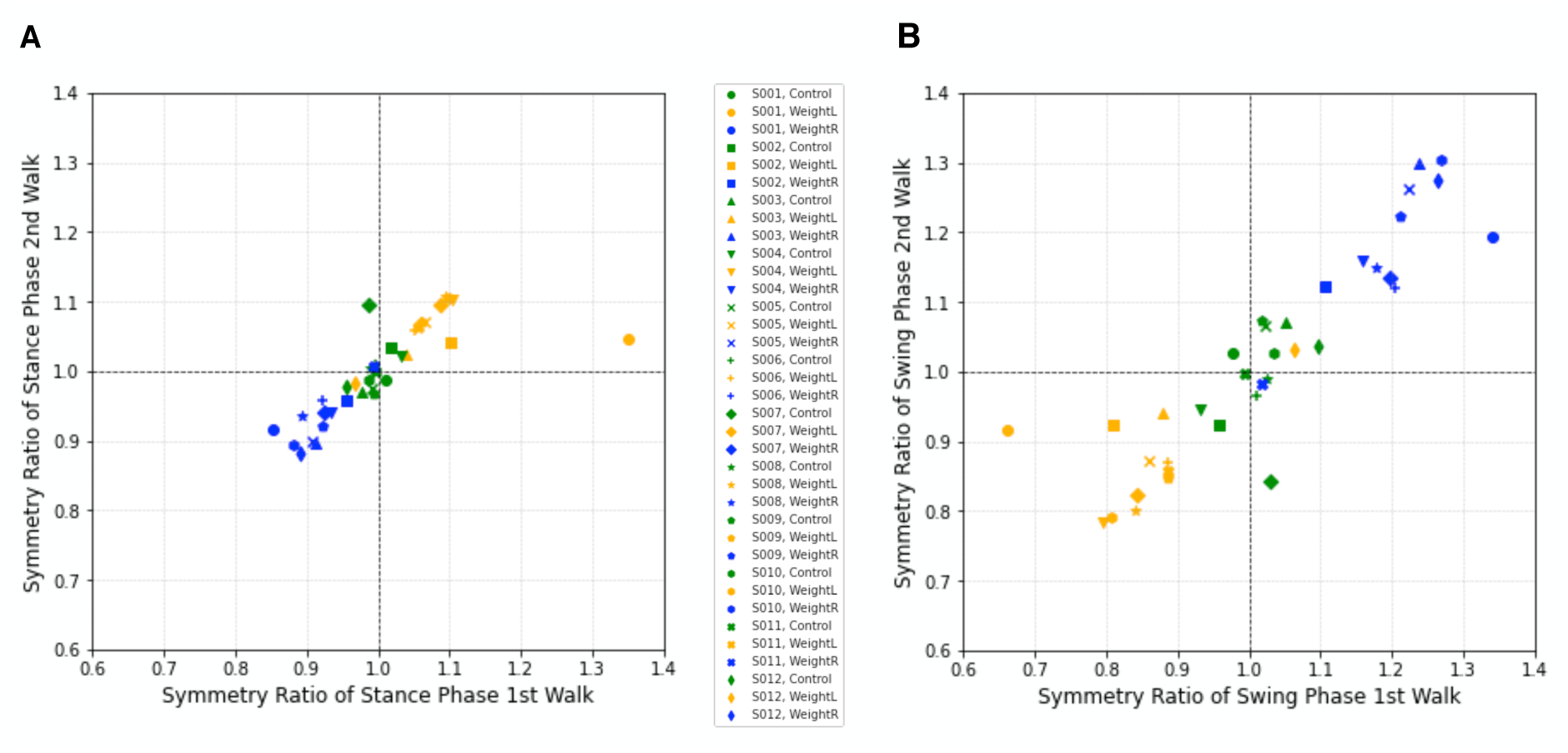
Validation on healthy subjects of the capability of the employed system (sensors plus computational pipeline) to measure gait asymmetry (asymmetry validation study, AVS). **(A)** Symmetry ratio of stance phase for healthy subjects of AVS, walking with weights on the left ankle (gold), right ankle (blue) or with no weight (green). **(B)** Symmetry ratio of swing phase for healthy subjects of AVS, walking with weights on the left ankle (gold), right ankle (blue) or no weight (green). Symmetry ratios for gait cycle duration and cadence are reported in Fig. S14. AVS is described in Methods Sec. 4.1, and we provided extended details in [43]. These results are summarized in Sec. 4.6.

**Figure S14:**
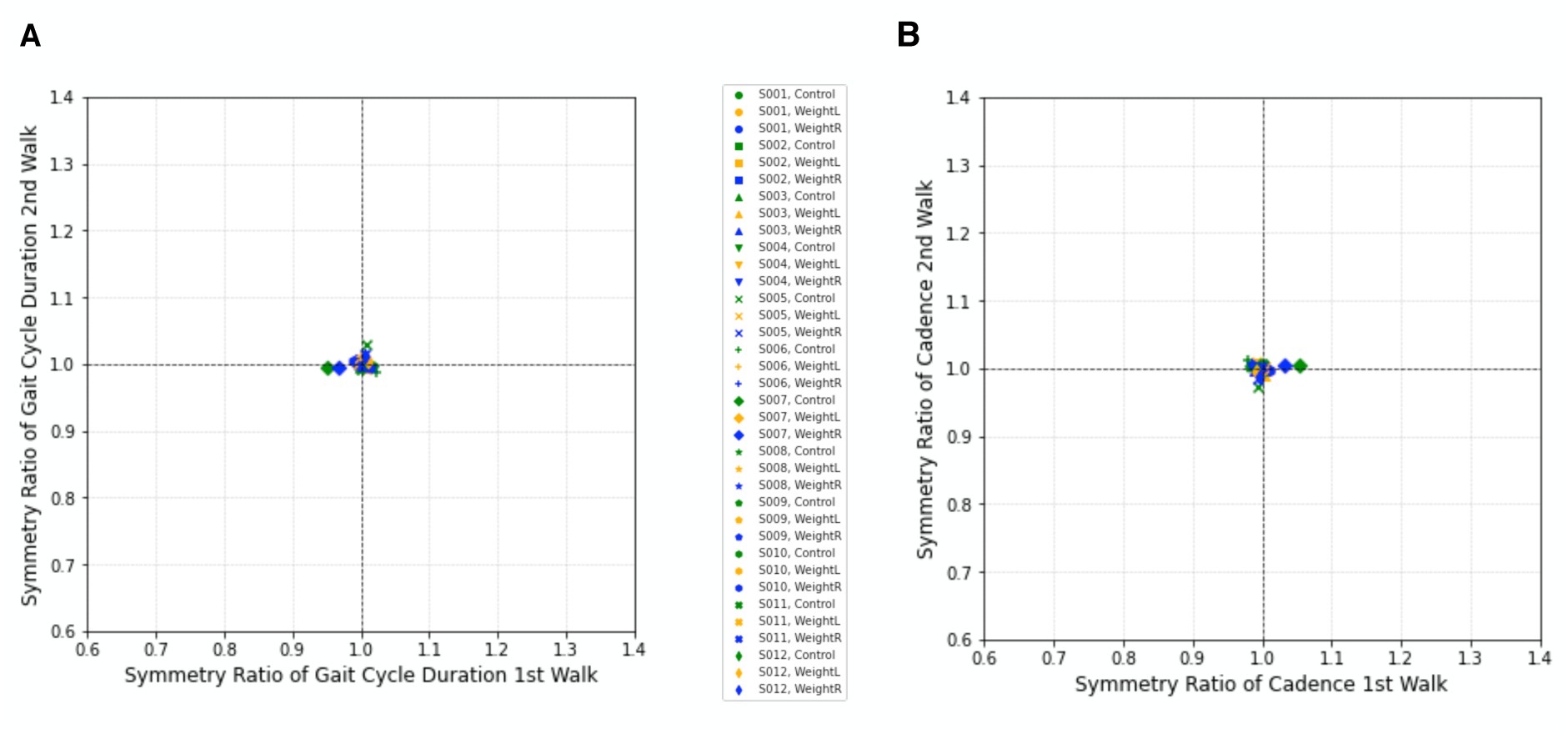
Symmetry ratios for gait cycle duration and cadence, from the asymmetry validation study (AVS) on healthy subjects. **(A)** Symmetry ratio of gait cycle duration for healthy subjects of AVS, walking with weights on the left ankle (gold), right ankle (blue) or no weight (green). **(B)** Symmetry ratio of cadence for healthy subjects of AVS, walking with weights on the left ankle (gold), right ankle (blue) or no weight (green). AVS is described in Methods Sec. 4.1, and we provided extended details in [43].

**Table S1:** Details of PD patients for clinical study (CS), see Methods Sec. 4.1.

| Counter | Disease | Conditions of examination | Gender | PD type | Clinical manifestation |
| --- | --- | --- | --- | --- | --- |
| 1 | PD | LCT (no DBS)* | M | NA | TD |
| 2 | PD | LCT (no DBS)* | M | NA | RD |
| 3 | PD | LCT (no DBS)* | F | IPS | RD |
| 4 | PD | LCT (no DBS)* | M | IPS | TD |
| 5 | PD | LCT (no DBS)* | M | IPS | EQ |
| 6 | PD | LCT (no DBS)* | M | NA | TD |
| 7 | PD | DBS and LDOPA off** | M | IPS | TD |
| 8 | PD | LCT (no DBS)* | M | IPS | TD |
| 9 | PD | DBS and LDOPA off** | M | IPS | RD |
| 10 | PD | LCT (no DBS)* | M | IPS | EQ |
| 11 | PD | LCT (no DBS)* | F | IPS | RD |
| 12 | PD | LCT (no DBS)* | M | IPS | RD |

\*LCT, levodopa challenge test. Measurement with withdrawn LDOPA more than 12 hours.
\*\*DBS off. LDOPA withdrawn more than 12 hours.
IPS, Idiopathic parkinson syndrome
TD, tremor dominant; RD, rigor dominant; EQ, equivalence type

**Table S2:** Details of NPH patients for clinical study (CS), see Methods Sec. 4.1.

| Counter | Disease | Conditions of examination | Gender |
| --- | --- | --- | --- |
| 1 | NPH | before LP | F |
| 2 | NPH | before LP | M |
| 3 | NPH | before LP | M |
| 4 | NPH | before LP | M |
| 5 | NPH | before LP | M |
| 6 | NPH | before LP | M |
| 7 | NPH | before LP | M |
| 8 | NPH | before LP | F |
| 9 | NPH | before LP | F |
| 10 | NPH | before LP | F |
| 11 | NPH | before LP | F |

## Notes

### Competing Interest Statement

The authors have declared no competing interest.

### Author Declarations

The clinical study contained in this work was conducted at the Centre Hospitalier de Luxembourg according to the principles of the Declaration of Helsinki (2013). The Ministry of Health of Luxembourg and the National Ethics Board (Comite National d'Ethique de Recherche, CNER) gave ethical approval for the clinical study contained in this work (Reference number: 202101/02, 0622-2). The validation study contained in this work was carried out on healthy subjects at the Trier University of Applied Sciences according to the principles of the Declaration of Helsinki (2013). The Ethics committee of the State Chamber of Medicine in Rhineland-Palatinate, Germany, gave ethical approval for the validation study contained in this work (Reference name: VitalMove Study).

